# Exposure to heavy metals *in utero* and autism spectrum disorder at age 3: A meta-analysis of two longitudinal cohorts of siblings of children with autism

**DOI:** 10.1101/2023.11.21.23298827

**Authors:** John F. Dou, Rebecca J. Schmidt, Heather E. Volk, Manon M. Nitta, Jason I. Feinberg, Craig J. Newschaffer, Lisa A. Croen, Irva Hertz-Picciotto, M. Daniele Fallin, Kelly M. Bakulski

**Affiliations:** University of Michigan, Ann Arbor, Michigan, USA; University of California Davis, Davis, California, USA; Johns Hopkins University, Baltimore, Maryland, USA; Penn State University, State College, PA, USA; Division of Research, Kaiser Permanente Northern California, Oakland, CA, USA; Rollins School of Public Health, Emory University, Atlanta, GA, USA

**Keywords:** Metals exposure, autism spectrum disorder, pregnancy cohort, epidemiology, cadmium

## Abstract

**Background:** Autism spectrum disorder (ASD) is a prevalent and heterogeneous neurodevelopmental disorder. Risk is attributed to genetic and prenatal environmental factors, though the environmental agents are incompletely characterized.

**Methods:** In Early Autism Risk Longitudinal Investigation (EARLI) and Markers of Autism Risk in Babies Learning Early Signs (MARBLES), two pregnancy cohorts of siblings of children with ASD, maternal urinary metals concentrations at two time points during pregnancy were measured using inductively coupled plasma mass spectrometry. At age three, clinicians assessed ASD with DSM-5 criteria. Using multivariable log binomial regression, we examined each metal for association with ASD status, adjusting for gestational age at urine sampling, child sex, maternal age, and maternal education, and meta-analyzed across the two cohorts.

**Results:** In EARLI (n=170) 17.6% of children were diagnosed with ASD, and an additional 43.5% were classified as having other non-neurotypical development (Non-TD). In MARBLES (n=156), 22.7% were diagnosed with ASD, while an additional 11.5% had Non-TD. In earlier pregnancy metals measures, having cadmium concentration over the level of detection was associated with 1.78 (1.19, 2.67) times higher risk of ASD, and 1.43 (1.06, 1.92) times higher risk of Non-TD. A doubling of early pregnancy cesium concentration was marginally associated with 1.81 (0.95, 3.42) times higher risk of ASD, and 1.58 (0.95, 2.63) times higher risk of Non-TD.

**Conclusion:** Exposure *in utero* to elevated levels of cadmium and cesium, as measured in maternal urine collected during pregnancy, was associated with increased risk of developing ASD.

## Introduction

Autism Spectrum Disorder (ASD) presents a major public health concern. ASD is a neurodevelopmental disorder characterized by impairments in social communication, social interaction, and restrictive and repetitive behavioral patterns and interests (Diagnostic and statistical manual of mental disordersL: DSM-5. 2013). In the United States, 1 in 36 children are affected by ASD, with the prevalence among boys 3.8 times greater than among girls (Maenner et al. 2023). Individuals with ASD and their families face significant social and financial burdens, with higher costs for individuals with more severe ASD (Rogge and Janssen 2019). The social cost of ASD was greater than $7 trillion between the years 1990 – 2019 and is projected to be an additional $4 to $15 trillion by 2029 (Cakir et al. 2020). Increases in ASD prevalence have been attributed to increasing social awareness (Keyes et al. 2012), changes to diagnostic criteria (Hansen et al. 2015), and participation in early intervention services (Worley et al. 2011). However, the full source of this increase is largely unknown, suggesting incidence may be rising. Environmental exposures may play a role in this increase. Understanding modifiable risk factors for ASD could play a major role in guiding public health interventions.

Metals are a potential modifiable risk factor in ASD. In the United States, women of childbearing age experience widespread environmental exposure to metals, and higher concentrations have been observed in pregnant woman compared to non-pregnant women (Martin and Fry 2018; Watson et al. 2020). Important neurodevelopmental processes occur during pregnancy (Estes and McAllister 2016), and exposure to environmental factors such as metals are suggested to have a role in ASD etiology (Bölte et al. 2019; Heyer and Meredith 2017; Lyall et al. 2014). Among children diagnosed with ASD relative to controls, higher blood levels of arsenic (Ding et al. 2023), mercury (Ding et al. 2023; Jafari et al. 2017; Zhang et al. 2021), lead (Rashaid et al. 2021; Saghazadeh and Rezaei 2017; Zhang et al. 2021), and cadmium (Baj et al. 2021) have been observed. Although these findings are suggestive, exposure to metals was measured after ASD diagnosis. In studies where child levels of metals were measured after ASD was diagnosed, it is not known if elevated exposure levels preceded ASD. Some studies have examined exposure during pregnancy. Poorer performance on social and behavioral tests among children at age 3 was associated with elevated manganese levels in infant toenails and arsenic in maternal toenails (Doherty et al. 2020), and prenatal maternal blood lead levels (Fruh et al. 2019). In contrast, elevated copper levels in maternal urine or blood during pregnancy was associated with decreased behavior problems assessed in children aged 3-7 years (Jedynak et al. 2021). Studies examining ASD diagnosis and metals exposure during pregnancy are less common. One nested case-control study in the Norwegian Mother, Father, and Child Cohort Study linked with the Norwegian Patient Registry examined maternal blood metals concentrations during pregnancy, finding elevated arsenic, cadmium, and manganese were associated with ASD, and lower levels of cesium, copper, mercury, and zinc were associated with ASD (Skogheim et al. 2021). There is limited study on prenatal metals exposure and ASD, and more prospective cohorts with exposure measures of multiple metals are needed.

This study was conducted in two pregnancy cohorts of siblings of children with ASD, the Early Autism Risk Longitudinal Investigation (EARLI) and the Markers of Autism Risk in Babies – Learning Early Signs (MARBLES) study. The goal of this study was to examine the associations between concentrations of a panel of twenty-two metals measured during pregnancy with ASD diagnosis in children at age 3 years.

## Methods

### Study sample

The Early Autism Risk Longitudinal Investigation (EARLI) is a prospective pregnancy cohort to study autism etiology (Newschaffer et al. 2012). The EARLI study was reviewed and approved by Human Subjects Institutional Review Boards (IRBs) from each of the four study sites (Johns Hopkins University, Drexel University, University of California Davis, and Kaiser Permanente Northern California). Markers of Autism Risk Learning Early Signs (MARBLES) is also an enriched-familial risk prospective pregnancy cohort to study autism etiology (Hertz-Picciotto et al.). The MARBLES protocol was reviewed and approved by the Human Subjects IRB from University of California Davis. Secondary data analysis for this manuscript was approved by the Human Subjects IRB for the University of Michigan. These studies recruited mothers of children with clinically confirmed ASD (probands) who were early in a subsequent pregnancy or were trying to become pregnant. Siblings of children with ASD are more likely to have a diagnosis of ASD or other developmental delays (Hansen et al. 2019; Miller et al. 2019). In EARLI there were 232 mothers with a subsequent child (sibling) born during the study between November 2009 and March 2012. In MARBLES there were 389 enrolled mothers that gave birth to 425 subsequent children (sibling) between December 1, 2006 and July 1, 2016.

### Covariate and outcome assessment

Demographics, pregnancy behaviors, and medical history were all collected via maternal questionnaire at enrollment. For the children born during the study (siblings), at age three years clinicians assessed them with DSM-5 criteria. Children were categorized into three groups: typically developing, ASD, or other non-typical development. Outcome categorization, based on a previously published algorithm using the Autism Diagnostic Observation Schedule (ADOS) and the Mullen Scales of Early Learning (MSEL) (Ozonoff et al. 2014), has been described in these cohorts previously (Mordaunt et al. 2019; Philippat et al. 2018). In brief, the typical development group did not meet diagnostic criteria for ASD. Those that did not meet diagnostic criteria, but had ADOS scores within three points of the cutoff or MSEL scores 1.5 to 2 standard deviations below average, were categorized in the non-typical development group. Finally, those who met diagnostic DSM-5 criteria and ADOS scores over the cutoff were categorized in the ASD group.

### Exposure assessment

Maternal urine samples collected at two time points during pregnancy (earliest timepoint approximately mean 19 weeks of pregnancy, latest timepoint approximately mean 32 weeks pregnancy) had urinary concentrations of a panel of metals measured using inductively coupled plasma mass spectrometry by NSF International (Centers for Disease Control and Prevention method 3018.3, with modifications for the expanded metals panel and the Thermo Scientific iCAP RQ instrument). Metals measured include antimony, arsenic, barium, beryllium, cadmium, cesium, chromium, cobalt, copper, lead, manganese, mercury, molybdenum, nickel, platinum, selenium, thallium, tin, tungsten, uranium, vanadium, and zinc. Samples for both cohorts were randomized together into two laboratory runs and runs had variable limits of detection (LOD) (**Supplemental Table 1**). For example, for lead the LOD was either 0.1 ppb (83 samples in EARLI, 36 samples in MARBLES) or 0.2 ppb (262 samples in EARLI, 369 samples in MARBLES), depending on batch. To assess urinary dilution, specific gravity was measured by NSF International using an ATAGO handheld digital refractometer model PAL-10S. After excluding samples involved in a multiple birth (n=23), related siblings from non-multiple births (selecting one randomly to keep, n=18 samples dropped), or missing gestational age at collection (n=3), EARLI had 165 mothers with metals measures from two timepoints, and 15 mothers with a measure from one timepoint. MARBLES had 154 mothers with metals measures from two timepoints, and 97 mothers with a measure from one timepoint. Distribution of gestational age at urine sample collection are shown in **Supplemental Figure 1**.

As a sensitivity analysis, maternal blood concentrations during pregnancy of cadmium, manganese, lead, selenium, and total mercury were also measured in EARLI. Maternal venous blood samples were collected in trace metal free EDTA tubes. Blood samples from the first study visit (n=215) were used. Metal concentrations in maternal blood samples were measured by inductively coupled dynamic reaction cell plasma mass spectrometry by the US Centers for Disease Control and Prevention (ELAN DRC II, PerkinElmer Norwalk, CT) (method DLS 3016.8, Centers for Disease Control and Prevention). Micro-clotting of the archived blood prevented measures in half of samples, leaving n=104 with measured concentrations.

Data used in this manuscript is publicly available through the National Institute of Mental Health Data Archive (EARLI cohort repository: 1600, MARBLES cohort repository: 1946, EARLI/MARBLES metals repository: 2462) and through data requests to the Principal Investigators of cohorts (EARLI: MDF, MARBLES: RJS).

### Statistical analyses

We used R statistical software (version 4.0.2) for statistical analysis. Code to produce analyses is available (https://github.com/bakulskilab). Metals with less than approximately 10% of samples above the LOD were dropped from analysis (beryllium, platinum, tungsten, uranium, vanadium). Metals with less than 75% of samples above the LOD (antimony, cadmium, chromium, lead) were treated as binary variables, based on whether a sample was above or below the LOD. For the rest of the metals, concentrations were used as continuous variables.

We substituted all urinary metals measures quantitated with values below the LOD with the value of the LOD/square root of two (Hornung and Reed 1990). Metal concentrations were adjusted for specific gravity by multiplying concentrations by the ratio of [the median specific gravity – 1] and [sample specific gravity – 1] (Middleton et al. 2019). We then log_2_ transformed the continuous concentrations. Outlier metals concentrations >5 standard deviations from the mean were dropped from analyses. The number of samples dropped per metal are listed in **Supplemental Table 2**.

We separated urinary measures into earlier and later pregnancy timepoints. For those with two measures, the sample with lowest gestational age at collection was put into the earlier timepoint sample. For those with only one sample, gestational age at collection >= 28 weeks (third trimester) was considered late pregnancy, and otherwise samples were considered earlier pregnancy. We compared exposure levels in early and late pregnancy with Spearman correlation tests.

We calculated univariate descriptive statistics on each cohort using mean and standard deviation for continuous variables and count and frequency for categorical variables. The distributions of metal concentrations were described using mean, median, standard deviation, interquartile range, and the number and percent above the limit of detection. We calculated Spearman correlation of metals concentrations within each cohort. Separately for each cohort, we compared the bivariate sample characteristics by neurodevelopmental outcome (ASD, non-typically developing, typically developing) using ANOVA tests for continuous variables and chi-square tests for categorical variables.

To estimate the adjusted associations between urinary metals concentration in pregnancy and neurodevelopmental status, we used log binomial models to get estimates of relative risk. Due to convergence issues, we used the delta-method normal approximations for fitting models using the epitools package (Muller and MacLehose 2014). We estimated the association of each metal with ASD status relative to the typically developing group using the log_2_ transformed concentrations, adjusting for gestational age at urine sampling, child sex, maternal age, and maternal education. We also tested metals associations with non-typically developing status (typically developing as reference) in separate log binomial models.

Models were fit separately for each cohort, then meta analyzed together using the inverse variance method in the R meta package. We reported risk ratios (RR) and 95% confidence intervals (95% CI) for each association and visualized the results using forest plots. For metals that were modeled continuously, since concentrations were log-transformed, the reported associations are for a doubling in concentration. For metals that were modeled as binary, we reported the risk ratio for above versus below the limit of detection. To account for multiple comparisons, we also reported false discovery rate adjusted p-values.

We performed several sensitivity analyses to assess the robustness of our findings. Since runs for metals measures had variable limits of detection, which impacts binary categorization and imputation for values below limit of detection, we ran models adjusted for batch. We also performed multivariable logistic regression for each of our models to generate adjusted odds ratios (OR) that may be compared to the risk ratios and to prior findings in the literature. Lastly, we performed analyses on the subset of EARLI samples with maternal blood metals measures available and compared the findings to the findings in urinary metals.

## Results

### Sample descriptive statistics

At the earliest pregnancy timepoint, urinary metal concentrations were above the limit of detection in greater than 75% of the samples for 13 metals in each cohort (arsenic, barium, cesium, cobalt, copper, manganese, mercury, molybdenum, nickel, selenium, thallium, tin, and zinc) (**Supplemental Table 3**). In both EARLI and MARBLES, cobalt (Co) and nickel (Ni) concentrations had the strongest correlation (Spearman r=0.57 in EARLI, r=0.59 in MARBLES) (**Figure 1**).

**Figure 1.**
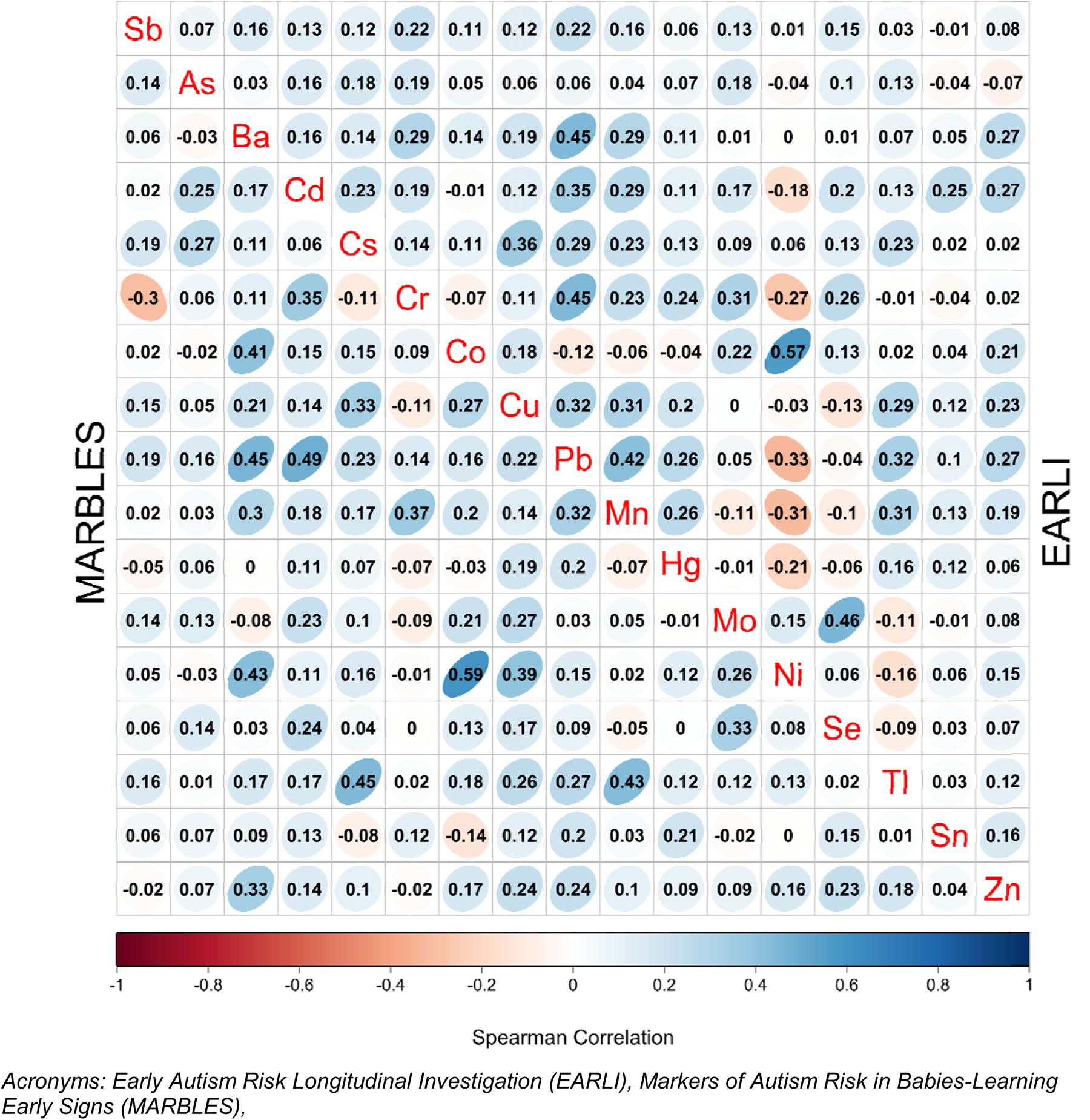
Spearman correlations of urinary metals concentrations, measured during early pregnancy, stratified by cohort. The upper right triangle shows the EARLI cohort. The lower left triangle shows the MARBLES cohort. Metals are represented by their chemical symbol along the diagonal.

In the earlier pregnancy timepoint there were metals concentrations available from 170 urine samples in EARLI (66 typically developing, 74 non-typically developing, 30 ASD) and 158 in MARBLES (104 typically developing, 18 non-typically developing, 36 ASD) (**Table 1**). In EARLI, maternal education and child sex assigned at birth were associated with child neurodevelopmental status. The typically developing group had higher levels of maternal education (71.2% with college degree), compared to the non-typically developing (54.1%) and ASD groups (43.3%). The typically developing and non-typically developing groups had a similar proportion of males (42.4% and 47.3%) but lower proportion than the ASD group (76.7%). In MARBLES, compared to the typically developing group (51.9% male), both the non-typically developing group (66.7% male) and ASD (75.0% male) had higher proportion of males (**Table 1**).

**Table 1.** Maternal and child characteristics of participants in the primary analytic sample with measures of metal exposure in urine from early pregnancy. Data are split by cohort and comparted by neurodevelopmental status of the sibling. Distributions of categorical variables are compared with a chi-square test and continuous variables are compared with ANOVA test.

| **EARLI cohort** | **Typically**<br>**developing**<br>**N=66** | **Non-typically**<br>**developing**<br>**N=74** | **Autism spectrum**<br>**disorder**<br>**N=30** | **P-value** |
| --- | --- | --- | --- | --- |
| Maternal Education |  |  |  | 0.020 |
| College Degree | 47 (71.2%) | 40 (54.1%) | 13 (43.3%) |  |
| No Degree | 19 (28.8%) | 34 (45.9%) | 17 (56.7%) |  |
| Maternal Age | 35.1 (4.68) | 33.3 (4.52) | 33.8 (3.87) | 0.057 |
| Infant Sex |  |  |  | 0.006 |
| F | 38 (57.6%) | 39 (52.7%) | 7 (23.3%) |  |
| M | 28 (42.4%) | 35 (47.3%) | 23 (76.7%) |  |
| Weeks of Gestation at<br>Sample Collection | 19.1 (5.14) | 18.3 (6.44) | 19.4 (7.76) | 0.607 |
| Infant Gestational Age<br>at Birth | 39.5 (1.40) | 39.5 (1.36) | 38.9 (2.00) | 0.094 |
| **MARBLES cohort** | **Typically**<br>**developing**<br>**N=104** | **Non-typically**<br>**developing**<br>**N=18** | **Autism spectrum**<br>**disorder**<br>**N=36** | **P-value** |
| Maternal Education |  |  |  | 0.259 |
| College Degree | 62 (59.6%) | 9 (50.0%) | 16 (44.4%) |  |
| No Degree | 42 (40.4%) | 9 (50.0%) | 20 (55.6%) |  |
| Maternal Age | 34.6 (4.73) | 33.8 (4.71) | 34.6 (5.01) | 0.822 |
| Infant Sex |  |  |  | 0.041 |
| Female | 50 (48.1%) | 6 (33.3%) | 9 (25.0%) |  |
| Male | 54 (51.9%) | 12 (66.7%) | 27 (75.0%) |  |
| Weeks of Gestation at<br>Sample Collection | 19.3 (3.99) | 18.7 (3.99) | 19.0 (3.98) | 0.803 |
| Infant Gestational Age<br>at Birth | 38.9 (1.39) | 38.9 (1.54) | 39.3 (1.04) | 0.296 |

*Acronyms: Early Autism Risk Longitudinal Investigation (EARLI), Markers of Autism Risk in Babies-Learning Early Signs (MARBLES)*

At the later pregnancy timepoint, urinary metal concentrations were above the limit of detection in slightly less than 75% of the sample for manganese, mercury, and tin (**Supplemental Table 4**), however they were modelled as continuous for comparability with the early timepoint results. In late pregnancy, cobalt and nickel remained the strongest correlated metals in MARBLES (Spearman r=0.74), but not in EARLI. In both cohorts, lead and copper (r=0.43 in EARLI, r=0.49in MARBLES) as well as lead and manganese (r=0.45 in both) were correlated (**Supplemental Figure 2**).

At the later pregnancy timepoint, there were 171 samples with urinary metal concentrations available in EARLI (65 typically developing, 75 non-typically developing, 31 ASD) and 231 in MARBLES (146 typically developing, 34 non-typically developing, 51 ASD) (**Supplemental Table 5**). For mothers with two timepoints, correlation between the two were strongest for measured cadmium (r=0.48 in EARLI, r=0.42 in MARBLES), cesium (r=0.42 in both cohorts), mercury (r=0.48 in EARLI, r=0.63 in MARBLES), tin (r=0.57 in EARLI, r=0.59 in MARBLES), and zinc (r=0.53 in EARLI, r=0.47 in MARBLES). Cross timepoint correlation was weakest for antimony (r=0.29 in EARLI, r=0.15 in MARBLES) and manganese (r=0.21 in EARLI, r=0.12 in MARBLES) (**Supplemental Table 6**).

### Urinary Metal Association with Autism Spectrum Disorder Status

We examined associations between urinary metals in the earlier pregnancy timepoint and ASD. In meta-analysis, comparing ASD to typical development, having urine cadmium concentration above the limit of detection was associated with RR=1.78 (95% CI 1.19, 2.67) times higher risk for ASD (EARLI RR=1.92, 95% CI 1.05, 3.51; MARBLES RR=1.68, 95% CI 0.97, 2.89). (**Figure 2**, **Table 2**). A doubling in arsenic was associated with lower ASD risk (RR=0.85, 95% CI 0.76, 0.94), driven by the EARLI cohort (EARLI RR=0.82, 95% CI 0.73, 0.92; MARBLES RR=1.01, 95% CI 0.78, 1.31). Marginal associations were observed with cesium, where a doubling in urinary concentration was estimated to have RR=1.81 (95% CI 0.95, 3.42). Thallium concentration doubling was marginally associated with RR=1.16 (95% CI 1.08, 1.26), with stronger effect in MARBLES (RR=1.15, 95% CI 1.10, 1.32) than in EARLI (RR=1.00, 95% CI 0.68, 1.48). The associations for cadmium (FDR=0.085) and arsenic (FDR=0.051) reached FDR < 0.1 when adjusting for multiple comparisons. No associations were observed between the remaining urinary metal concentrations and ASD status at this early pregnancy time point.

**Figure 2.**
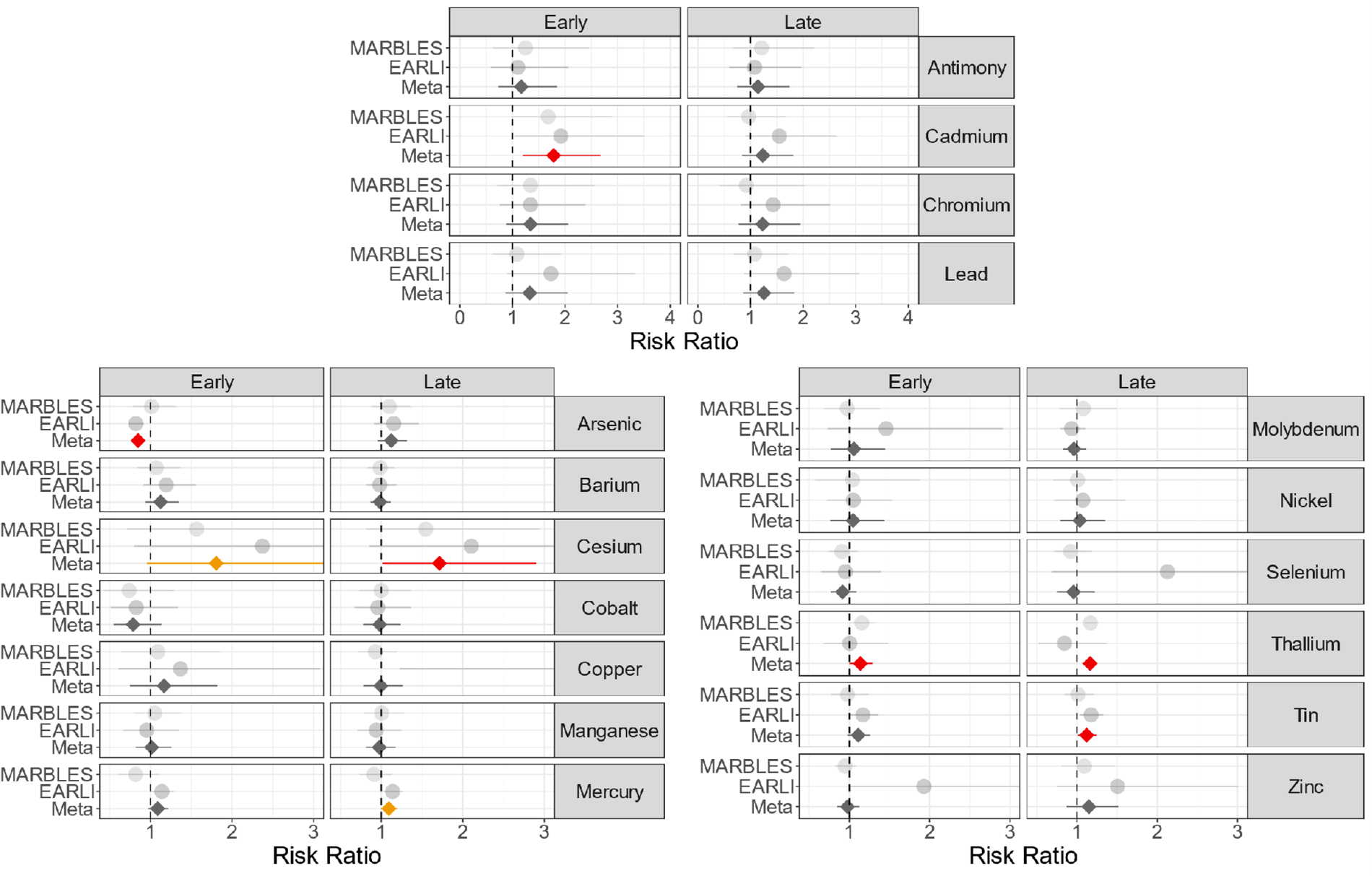
Adjusted risk ratios for the associations between maternal urinary metals concentrations measuring during pregnancy and risk of autism spectrum disorder, relative to typically developing. Antimony, cadmium, chromium, and lead compare over limit of detection vs under the limit of detection for that metal. Remaining metals show the risk ratio for a doubling in metal concentration. Analyses were performed stratified by cohort (EARLI and MARBLES) and then meta-analyzed across cohorts. Red denotes a nominal meta-analysis p-value < 0.05, and orange a nominal meta-analysis p-value < 0.10.

**Table 2.**
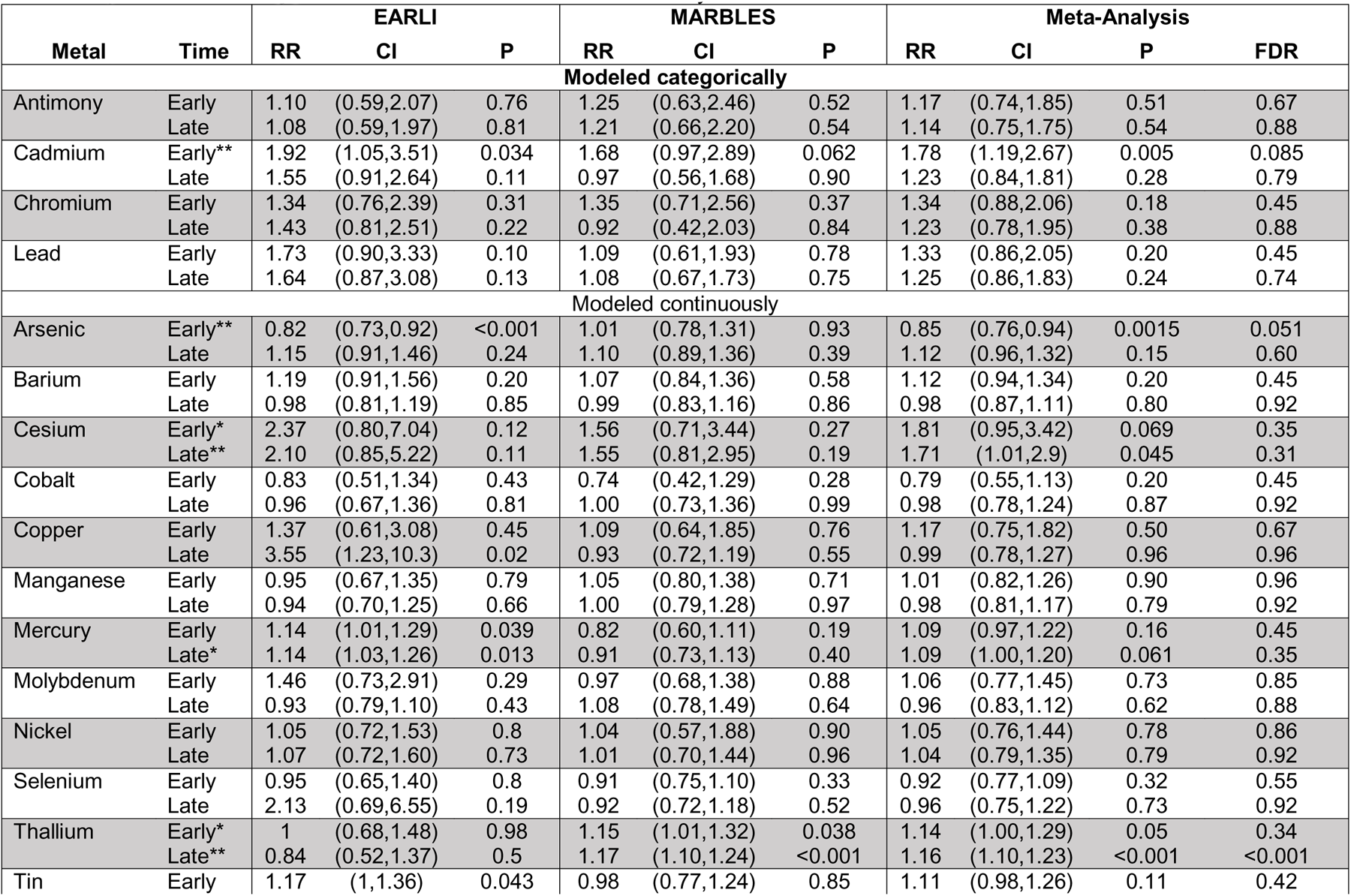
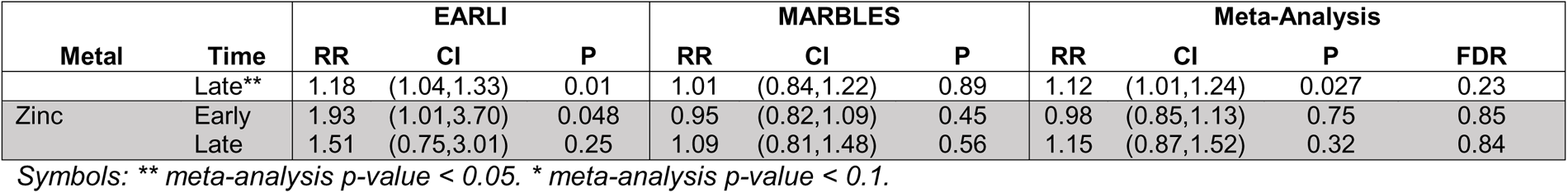
Adjusted risk ratios for the associations between maternal urinary metal concentrations measured during pregnancy and risk of autism spectrum disorder, relative to typically developing. Log binomial models were adjusted for gestational age, child sex, maternal age, and maternal education. Four metals were modeled categorically (above versus below the limit of detection) and the remaining metals were log_2_ transformed and modeled continuously.

At the later pregnancy timepoint, we estimated the association between each metal concentration and ASD. Comparing ASD to typically developing in meta-analyses, a doubling in cesium was associated with RR=1.71 (95% CI 1.01, 2.9) in meta-analysis (EARLI RR=2.10, 95% CI 0.85, 5.22.; MARBLES RR=1.55, 95% CI 0.81, 2.95) (**Figure 2**, **Table 2**). A doubling in thallium was associated with ASD with RR=1.16 (95% CI 1.10,1.23), though effects were different between cohorts (EARLI RR=0.84, 95% CI 0.52, 1.37.; MARBLES RR=1.17, 95% CI 1.10, 1.24). A doubling in tin was associated with RR=1.12 (95% CI 1.01,1.24). A doubling in mercury was marginally associated with RR=1.09 (95% CI 1.00, 1.20), with differences between EARLI (RR=1.14, 95% CI 1.03, 1.26) and MARBLES (RR=0.91, 95% CI 0.73, 1.13). The association with thallium reached FDR < 0.1. No associations were observed between the remaining urinary metal concentrations and ASD status at this later pregnancy time point.

### Urinary Metal Association with Non-Typically Developing Status

We repeated the adjusted regression analyses estimating the association of earlier pregnancy urinary metals and non-typically developing status. Urine cadmium concentrations above the LOD was associated with elevated risk of non-typical development, with RR=1.43 (95% CI 1.06, 1.92). (**Figure 3**, **Table 3**). A doubling of cesium urinary concentration was marginally associated with RR=1.58 (95% CI 0.95, 2.63) times higher risk of non-typical development. A marginal relationship with molybdenum was also observed, where a doubling in concentration was related to RR=1.47 (95% CI 0.95, 2.28). A doubling of nickel was associated with RR=1.40 (95% CI 1.01, 1.94), driven by the EARLI cohort with RR=1.55 (95% CI 1.08, 2.23). No associations were observed between the remaining urinary metal concentrations and non-typically developing status at this early pregnancy time point.

**Figure 3.**
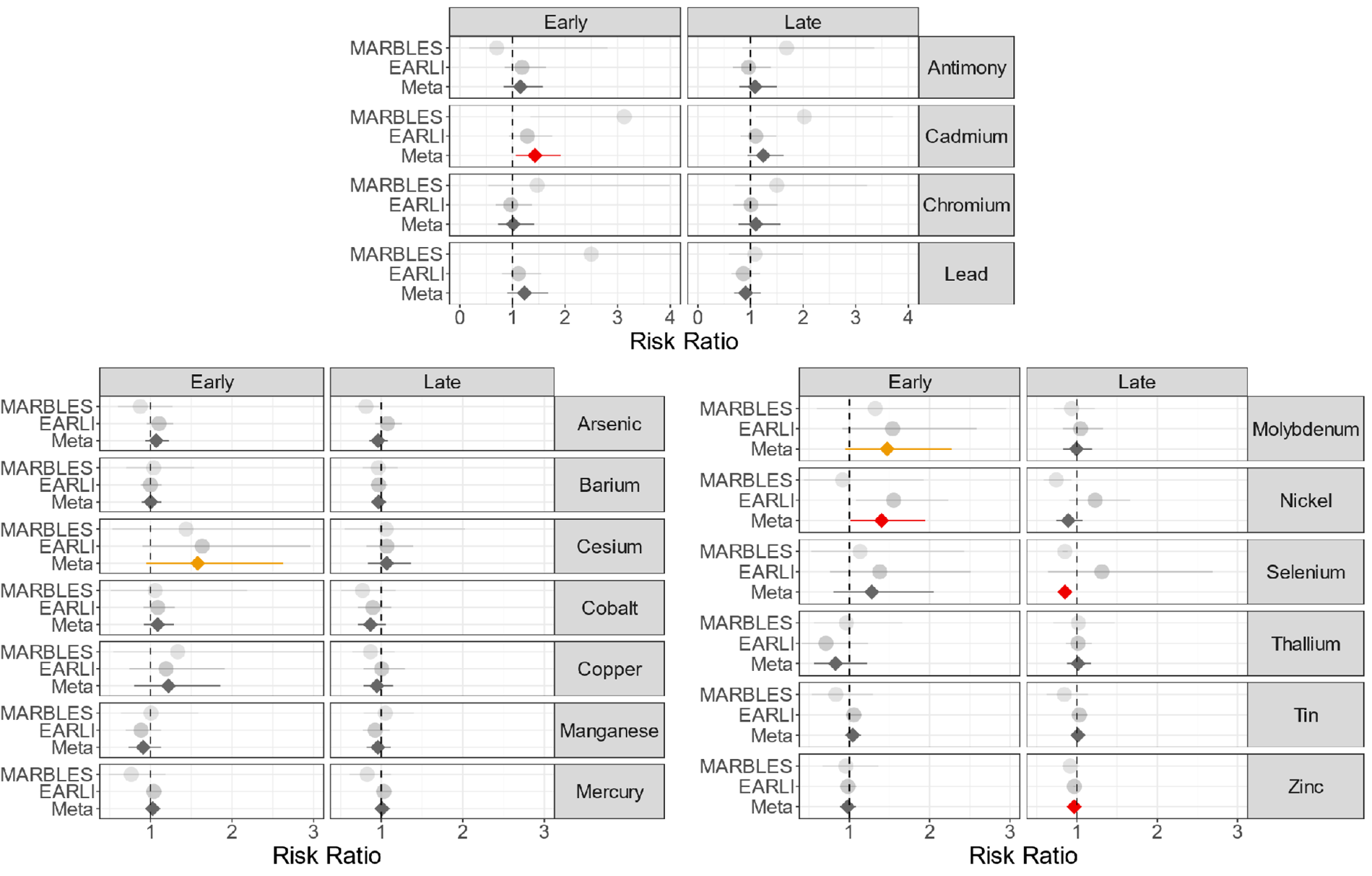
Adjusted risk ratios for the associations between maternal urinary metals concentrations measuring during pregnancy and risk of non-typically developing, relative to typically developing. Antimony, cadmium, chromium, and lead compare over limit of detection vs under the limit of detection for that metal. Remaining metals show risk ratio for a doubling in metal concentration. Analyses were performed stratified by cohort (EARLI and MARBLES) and then meta-analyzed across cohorts. Red denotes a nominal meta-analysis p-value < 0.05, and orange a nominal meta-analysis p-value < 0.10.

**Table 3.**
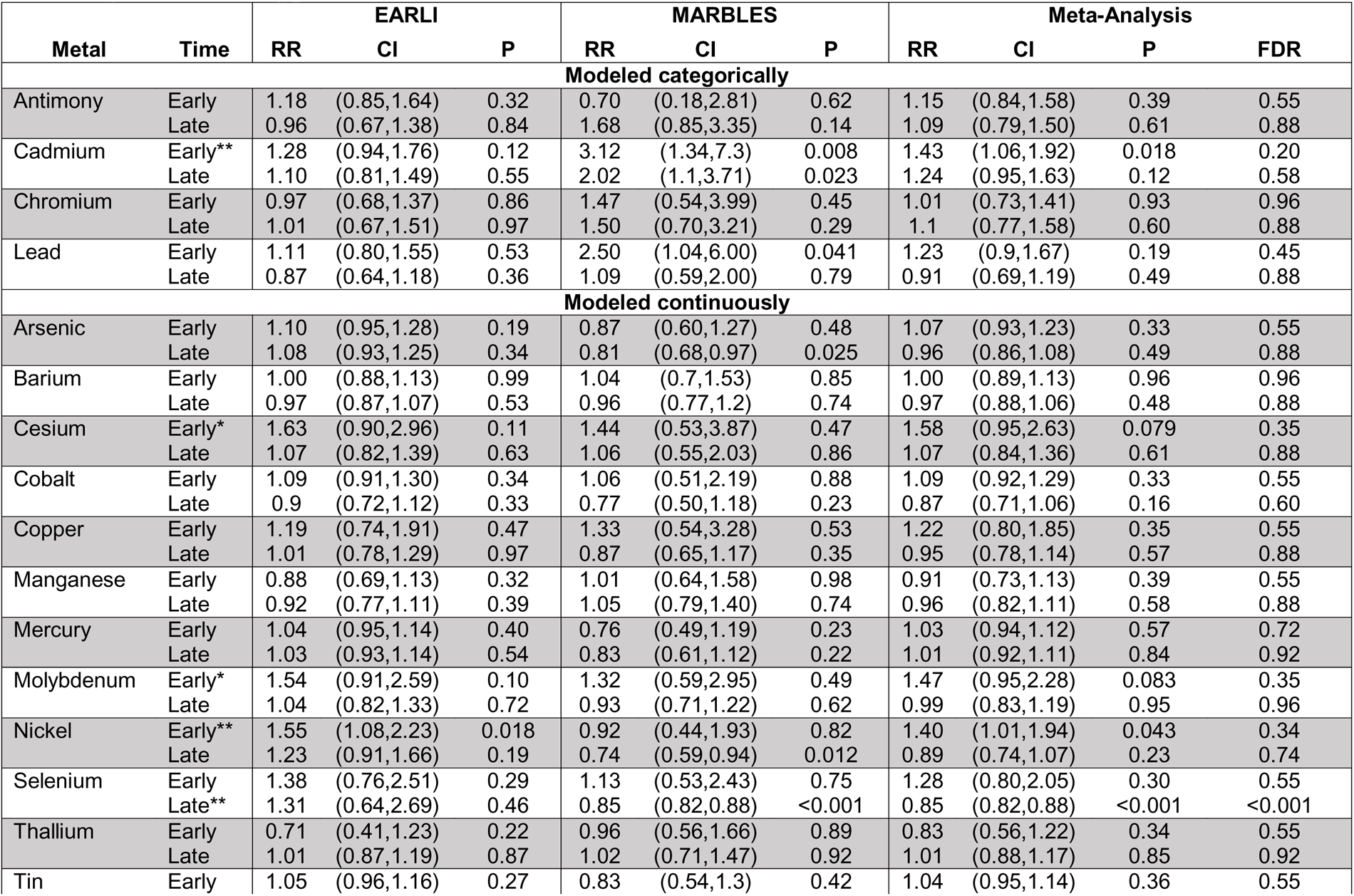
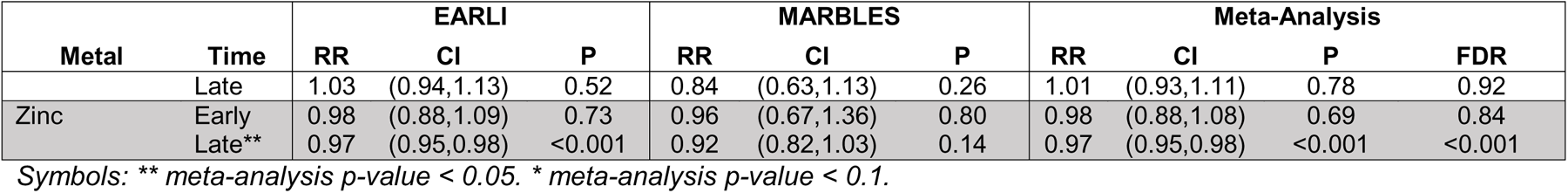
Adjusted risk ratios for the associations between maternal urinary metal concentrations measured during pregnancy and risk of non-typically developing, relative to typically developing. Log binomial models were adjusted for gestational age, child sex, maternal age, and maternal education. Four metals were modeled categorically (above versus below the limit of detection) and the remaining metals were log_2_ transformed and modeled continuously.

We examined associations between non-typically developing and the later pregnancy metals measures. Though not statistically significant, having urine cadmium concentration above the limit of detection was associated with RR=1.24 (95% CI 0.95, 1.63) times higher risk of non-typically developing status in meta-analysis (**Supplemental Figure 3, Table 3**). A doubling of the essential metal selenium concentration was associated in meta-analysis with RR=0.85 (95% CI 0.82, 0.88) times lower risk of non-typically developing status, driven by precision of results in MARBLES and had opposite directions of effect by cohort (EARLI RR=1.31, 95% CI 0.64, 2.69; MARBLES RR=0.85, 95% CI 0.82, 0.88). A doubling of the essential metal zinc concentration was associated with RR=0.97 (95% CI 0.95, 0.98) lower risk of non-typically developing status. The zinc and selenium associations reached FDR < 0.1. No associations were observed between the remaining urinary metal concentrations and non-typically developing status at this late pregnancy time point.

### Maternal Blood Metal Association with Neurodevelopmental Status

In EARLI, 92 maternal blood samples collected during pregnancy had available covariate and blood metals measures (41 typically developing, 32 non-typically developing, 19 ASD) (**Supplemental Table 7**). A doubling in maternal blood cadmium was marginally associated with RR=1.11 (95% CI 0.96, 1.29) higher risk of ASD, and a doubling in maternal blood lead was associated with RR=1.23 (95% CI 1.01, 1.54) higher risk of ASD (**Supplemental Figure 3**). A doubling in cadmium was also associated with RR=1.10 (95% CI 1.02, 1.19) higher risk of non-typical development. A doubling in maternal blood lead was associated with RR=1.16 (95% CI 1.00, 1.35) higher risk of non-typical development (**Supplemental Figure 2**). No associations were observed between the remaining blood metal concentrations (mercury, selenium, manganese) and neurodevelopmental status.

### Sensitivity Analysis

With batch as a covariate (**Supplemental Tables 8 and 9**), the cadmium association in early pregnancy with ASD remained consistent where being over the limit of detection was associated with RR=1.78 (95% CI 1.19, 2.65) times higher risk of ASD. The cadmium associations with non-typical development also remained consistent. With batch adjustment, antimony in earlier pregnancy was associated with ASD, with RR=1.58 (95% CI 1.01, 2.48). The associations between cesium and ASD as well as non-typical development remained consistent with slight attenuation, as did associations between earlier pregnancy molybdenum and nickel with non-typical development. On the other hand, the relationships with thallium and ASD were attenuated.

Using logistic regression models, consistency to the previous log binomial findings was observed for cadmium and cesium. In general, estimates on the odds ratio scale were higher in magnitude and significance for cadmium and cesium. Strength of relationships between arsenic, mercury, thallium, and tin with ASD were attenuated with larger confidence intervals when using logistic regression (**Supplemental Table 10**). Earlier pregnancy molybdenum and nickel associations with non-typical development remained consistent in logistic regression, while selenium and zinc associations with non-typical development were attenuated (**Supplemental Table 11**). In logistic regression, earlier pregnancy lead had stronger association with non-typical development, where lead over the LOD was marginally associated with OR=1.68 (95% CI 0.92, 3.07).

## Discussion

In these two prospective birth cohorts of siblings of children with ASD, we measured maternal urinary metals levels during two timepoints in pregnancy and examined relationships to ASD or non-typical development status at age 3. To our knowledge, this is the first study to report associations between maternal prenatal urine heavy metal concentrations and ASD diagnosis in children at 3 years of age in such cohorts. Our most consistent finding was heightened risk of atypical neurodevelopment related to cadmium exposure. Although the relationships were not significant in late pregnancy, the directions of effect were consistent across time periods. Furthermore, similar findings were observed in the maternal blood subsample. Cesium related to atypical neurodevelopment was also notable, with consistency across ASD and non-typical development outcomes and timepoints, with exception of later pregnancy cesium and non-typical development. Cadmium and cesium associations were also the most robust to different modelling strategies. This study suggests metals exposure during pregnancy may be related to risk of ASD or non-typical development status at age 3.

Existing studies have examined the relationship between heavy metals exposure and ASD with considerable heterogeneity in exposure timing and matrices measured (Campbell et al. 2021). A systematic review and meta-analysis of lead concentrations in children with ASD from cross-sectional and case-control studies showed significant difference in blood lead levels compared to controls, but not in urinary lead levels (Nakhaee et al. 2023). This mirrors our results in maternal measures during pregnancy, where we found maternal blood lead levels were associated with risk of ASD or non-typical development in offspring, but not maternal urinary lead levels. The study in the Norwegian Mother, Father, and Child Cohort Study found higher odds of ASD for children in the highest quartile of cadmium exposure measured in maternal blood during pregnancy (Skogheim et al. 2021), matching results from the present study. The same study found the highest quartile of maternal blood cesium levels had lower odds of ASD compared to the lowest quartile, while in contrast our study suggests higher risk of ASD with higher maternal urinary cesium. Our results for selenium were mixed, doubling of late pregnancy selenium concentration was associated with lower risk of non-typical development, however there were opposite effect estimates between cohorts. Selenium supplementation in an animal model attenuated autism phenotype (Wu et al. 2022a), and studies measuring selenium cross-sectionally in children in Saudi Arabia (El-Ansary et al. 2017) and China (Wu et al. 2022b) found lower selenium levels in those with ASD. On the other hand, two-sample Mendelian randomization analysis using genetic instruments of blood and blood-toenail selenium suggest selenium levels are associated with increased risk of ASD (Guo et al. 2023), and in the Boston Birth Cohort maternal red blood cell selenium levels measured at near delivery were associated with increased odds of ASD in children (Lee et al. 2021). Considerable heterogeneity in direction of association between selenium and ASD exist in our study and in the literature, along with heterogeneity in timing and tissue of measurements. Our findings add to a growing body of evidence of the neurodevelopmental impacts of metals exposure during pregnancy.

This study has several strengths. We were able to assay a wide array of metals with high detection rates in two different birth cohorts, at two different timepoints. In one cohort, we were also able to evaluate five metals in a different exposure matrix: maternal blood during pregnancy. The longitudinal design allowed examination of exposure measures during pregnancy that preceded subsequent ASD outcome 36 months after birth. The enriched risk cohort design ensured all participants were clinically assessed using gold standards for ASD diagnosis.

This study modeled metals as linear or dichotomous, but some metals, especially essential nutrients, may have non-linear relationships. While the sibling cohort design allowed for an extensively phenotyped sample, our findings may not be generalizable to populations where ASD is less common, thus it would be important to also compare to results found in population-based samples. Future studies should also consider other exposure matrices or timepoints. The choice of exposure matrix is also important for exposure timing. For example, blood cadmium levels reflect recent exposure, while urinary cadmium reflects a longer, cumulative exposure (Agency for Toxic Substances and Disease Registry (ATSDR) 2012). Certain exposure matrices may be more reliable for some metals. For example, blood lead is a more reliable measure of recent exposure compared to urinary or hair lead levels (Agency for Toxic Substances and Disease Registry (ATSDR) 2020), which may explain our findings of stronger blood lead ASD associations than those seen with urinary lead.

This study suggests that prenatal exposure to toxic metals, such as cadmium, impacts risk of ASD or non-typical development in offspring. Potential routes of exposure to metals include contamination of soil and water, through ambient air, and through use in industrial applications or domestic products (Tchounwou et al. 2012). Public health measures to reduce these exposures to heavy metals during pregnancy may be an important preventative strategy for neurodevelopmental disorders, though larger longitudinal studies are needed as well as studies to determine which routes of exposure are important for specific metals.

## Funding Acknowledgements

Funding for the EARLI study was provided by the National Institutes of Health (R01ES016443, R24ES030893) and Autism Speaks (003953). Funding for the MARBLES study was provided by the National Institutes of Health (R01ES020392, R01ES028089, R/U24ES028533, and P01ES011269) and the United States Environmental Protection Agency Science to Achieve Results program (#RD-83329201). Funding for metals measures and this work was supported by the National Institutes of Health (R01ES025531). The content is solely the responsibility of the authors and does not necessarily represent the official views of the National Institutes of Health.

