## Supplemental Material for "Exposure to heavy metals *in utero* and autism spectrum disorder at age 3: A meta-analysis of two longitudinal cohorts of siblings of children with autism"

**Supplementary Information**

This supplementary information contains 11 supplemental tables and 3 supplemental figures.

**Supplemental Table 1.** Number of samples by cohort, pregnancy timepoint, and batches having different limits of detection.

Separate file: Supplemental Table 1 Limits of Detection Descriptives.csv

**Supplemental Table 2**. Number of samples dropped from analysis based upon being an outlier > 5 standard deviations from the mean in log transformed measures.

| **Metal** | **EARLI Early Timepoint** | **EARLI Late Timepoint** | **MARBLES Early Timepoint** | **MARBLES Late Timepoint** |
| --- | --- | --- | --- | --- |
| Antimony | 0 | 0 | 0 | 1 |
| Arsenic | 0 | 0 | 0 | 0 |
| Barium | 0 | 0 | 0 | 0 |
| Cadmium | 0 | 0 | 0 | 0 |
| Cesium | 0 | 0 | 0 | 1 |
| Chromium | 0 | 0 | 0 | 1 |
| Cobalt | 0 | 0 | 0 | 0 |
| Copper | 0 | 1 | 0 | 3 |
| Lead | 0 | 0 | 0 | 1 |
| Manganese | 0 | 0 | 0 | 2 |
| Mercury | 0 | 0 | 0 | 0 |
| Molybdenum | 0 | 0 | 0 | 0 |
| Nickel | 0 | 0 | 0 | 1 |
| Selenium | 0 | 0 | 0 | 0 |
| Thallium | 0 | 0 | 0 | 0 |
| Tin | 0 | 0 | 0 | 0 |
| Zinc | 0 | 0 | 0 | 0 |

**Supplemental Table 3.** Distribution of metals concentrations measured in earlier pregnancy urine samples by cohort. All metals concentrations are measured in parts per billion (ppb).

| **Metal** | **Cohort** | **Mean (ppb)** | **Standard deviation** | **Median (ppb)** | **Interquartile range** | **N Above LOD** | **N** | **Percent Above LOD** |
| --- | --- | --- | --- | --- | --- | --- | --- | --- |
| Antimony | EARLI | 0.0442 | 0.0292 | 0.0386 | 0.0252 | 48 | 173 | 27.7 |
|  | MARBLES | 0.0222 | 0.0981 | 0.0356 | 0.0516 | 27 | 167 | 16.2 |
| Arsenic | EARLI | 14.9 | 31.7 | 5.97 | 8.63 | 172 | 173 | 99.4 |
|  | MARBLES | 9.86 | 11.1 | 6.5 | 5.08 | 167 | 167 | 100 |
| Barium | EARLI | 2.39 | 2.15 | 1.82 | 2.18 | 171 | 173 | 98.8 |
|  | MARBLES | 2.42 | 1.86 | 1.89 | 2.05 | 164 | 167 | 98.2 |
| Beryllium | EARLI | 0.0106 | 0.0755 | -0.00311 | 0.049 | 21 | 173 | 12.1 |
|  | MARBLES | -0.00597 | 0.0676 | 0.00171 | 0.0503 | 7 | 167 | 4.2 |
| Cadmium | EARLI | 0.118 | 0.132 | 0.113 | 0.135 | 94 | 173 | 54.3 |
|  | MARBLES | 0.0944 | 0.104 | 0.0832 | 0.107 | 56 | 167 | 33.5 |
| Cesium | EARLI | 3.75 | 1.41 | 3.45 | 1.58 | 173 | 173 | 100 |
|  | MARBLES | 4.84 | 1.75 | 4.43 | 1.74 | 167 | 167 | 100 |
| Chromium | EARLI | 0.355 | 0.493 | 0.124 | 0.64 | 49 | 173 | 28.3 |
|  | MARBLES | 0.55 | 0.662 | 0.413 | 0.604 | 29 | 167 | 17.4 |
| Cobalt | EARLI | 0.697 | 0.39 | 0.607 | 0.459 | 173 | 173 | 100 |
|  | MARBLES | 0.974 | 0.486 | 0.877 | 0.532 | 166 | 167 | 99.4 |
| Copper | EARLI | 8.51 | 3.83 | 8.02 | 3.53 | 158 | 173 | 91.3 |
|  | MARBLES | 9.5 | 4.6 | 8.4 | 5 | 142 | 167 | 85 |
| Lead | EARLI | 0.253 | 0.214 | 0.214 | 0.236 | 109 | 173 | 63 |
|  | MARBLES | 0.223 | 0.239 | 0.165 | 0.194 | 69 | 167 | 41.3 |
| Manganese | EARLI | 0.256 | 0.214 | 0.231 | 0.187 | 136 | 173 | 78.6 |
|  | MARBLES | 0.332 | 0.31 | 0.262 | 0.195 | 132 | 167 | 79 |
| Mercury | EARLI | 0.282 | 0.243 | 0.202 | 0.271 | 156 | 173 | 90.2 |
|  | MARBLES | 0.251 | 0.238 | 0.172 | 0.27 | 127 | 167 | 76 |
| Molybdenum | EARLI | 59 | 39.4 | 47.4 | 39.5 | 173 | 173 | 100 |
|  | MARBLES | 62.3 | 33.1 | 54 | 35.8 | 167 | 167 | 100 |
| Nickel | EARLI | 3.4 | 2.04 | 3.05 | 2.6 | 145 | 173 | 83.8 |
|  | MARBLES | 5.83 | 2.09 | 5.47 | 2.65 | 161 | 167 | 96.4 |
| Platinum | EARLI | -0.0192 | 0.0453 | -0.0113 | 0.0453 | 5 | 173 | 2.9 |
|  | MARBLES | 0.00874 | 0.023 | 0.00605 | 0.02 | 3 | 167 | 1.8 |
| Selenium | EARLI | 40.4 | 18.4 | 35.9 | 16.4 | 173 | 173 | 100 |
|  | MARBLES | 41.8 | 17.7 | 37.7 | 19.4 | 167 | 167 | 100 |
| Thallium | EARLI | 0.129 | 0.0602 | 0.118 | 0.0622 | 170 | 173 | 98.3 |
|  | MARBLES | 0.132 | 0.0596 | 0.12 | 0.0765 | 159 | 167 | 95.2 |
| Tin | EARLI | 0.584 | 0.815 | 0.339 | 0.467 | 136 | 173 | 78.6 |
|  | MARBLES | 0.582 | 0.686 | 0.346 | 0.42 | 129 | 167 | 77.2 |
| Tungsten | EARLI | 0.119 | 0.155 | 0.0841 | 0.107 | 18 | 173 | 10.4 |
|  | MARBLES | 0.158 | 0.217 | 0.114 | 0.152 | 23 | 167 | 13.8 |
| Uranium | EARLI | -0.00897 | 0.0183 | -0.0042 | 0.0179 | 10 | 173 | 5.8 |
|  | MARBLES | -0.00101 | 0.0283 | 0.000559 | 0.0135 | 16 | 167 | 9.6 |
| Vanadium | EARLI | 0.2 | 0.167 | 0.161 | 0.183 | 6 | 173 | 3.5 |
|  | MARBLES | 0.35 | 0.375 | 0.268 | 0.186 | 6 | 167 | 3.6 |
| Zinc | EARLI | 266 | 163 | 224 | 192 | 173 | 173 | 100 |
|  | MARBLES | 276 | 185 | 232 | 208 | 167 | 167 | 100 |

*Acronyms: Early Autism Risk Longitudinal Investigation (EARLI), Markers of Autism Risk in Babies-Learning Early Signs (MARBLES), limit of detection (LOD)*

**Supplemental Table 4.** Distribution of metals concentrations measured in later pregnancy urine samples by cohort. All metals concentrations are measured in parts per billion (ppb).

| **Metal** | **Cohort** | **Mean** | **Standard deviation** | **Median** | **Interquartile range** | **Above LOD** | **N** | **Percent Above LOD** |
| --- | --- | --- | --- | --- | --- | --- | --- | --- |
| Antimony | EARLI | 0.0421 | 0.0241 | 0.0383 | 0.0294 | 43 | 172 | 25 |
|  | MARBLES | 0.0351 | 0.0885 | 0.0338 | 0.0495 | 37 | 238 | 15.5 |
| Arsenic | EARLI | 13.7 | 25.2 | 5.34 | 8.08 | 172 | 172 | 100 |
|  | MARBLES | 11.3 | 11.8 | 6.85 | 8.88 | 237 | 238 | 99.6 |
| Barium | EARLI | 3.39 | 2.92 | 2.58 | 2.84 | 166 | 172 | 96.5 |
|  | MARBLES | 3.91 | 4.23 | 2.67 | 3.41 | 237 | 238 | 99.6 |
| Beryllium | EARLI | 0.0127 | 0.0747 | 0.00281 | 0.0402 | 17 | 172 | 9.9 |
|  | MARBLES | -0.00739 | 0.0626 | -0.00669 | 0.0532 | 11 | 238 | 4.6 |
| Cadmium | EARLI | 0.103 | 0.134 | 0.09 | 0.13 | 83 | 172 | 48.3 |
|  | MARBLES | 0.0693 | 0.111 | 0.0636 | 0.112 | 62 | 238 | 26.1 |
| Cesium | EARLI | 3.42 | 1.38 | 3.14 | 1.75 | 171 | 172 | 99.4 |
|  | MARBLES | 4.59 | 1.63 | 4.29 | 2.21 | 238 | 238 | 100 |
| Chromium | EARLI | 0.276 | 0.456 | 0.0995 | 0.253 | 31 | 172 | 18 |
|  | MARBLES | 0.494 | 0.622 | 0.287 | 0.59 | 31 | 238 | 13 |
| Cobalt | EARLI | 1.1 | 0.561 | 1.03 | 0.672 | 171 | 172 | 99.4 |
|  | MARBLES | 1.51 | 0.737 | 1.38 | 0.841 | 238 | 238 | 100 |
| Copper | EARLI | 11.2 | 6.47 | 9.89 | 5.37 | 164 | 172 | 95.3 |
|  | MARBLES | 13.7 | 13.8 | 11.2 | 7.87 | 203 | 238 | 85.3 |
| Lead | EARLI | 0.254 | 0.235 | 0.197 | 0.264 | 108 | 172 | 62.8 |
|  | MARBLES | 0.301 | 0.601 | 0.174 | 0.236 | 98 | 238 | 41.2 |
| Manganese | EARLI | 0.285 | 0.223 | 0.237 | 0.269 | 128 | 172 | 74.4 |
|  | MARBLES | 0.374 | 0.421 | 0.27 | 0.256 | 172 | 238 | 72.3 |
| Mercury | EARLI | 0.224 | 0.207 | 0.161 | 0.168 | 146 | 172 | 84.9 |
|  | MARBLES | 0.227 | 0.288 | 0.137 | 0.212 | 148 | 238 | 62.2 |
| Molybdenum | EARLI | 52.5 | 37.8 | 44.1 | 30.6 | 172 | 172 | 100 |
|  | MARBLES | 62.1 | 40 | 50 | 44.8 | 238 | 238 | 100 |
| Nickel | EARLI | 4.04 | 2.23 | 3.55 | 2.52 | 153 | 172 | 89 |
|  | MARBLES | 6.48 | 3.03 | 5.92 | 3.92 | 224 | 238 | 94.1 |
| Platinum | EARLI | -0.0117 | 0.0456 | -0.00447 | 0.032 | 3 | 172 | 1.7 |
|  | MARBLES | 0.00473 | 0.0245 | 0.00223 | 0.0203 | 1 | 238 | 0.4 |
| Selenium | EARLI | 36.9 | 15.5 | 33.1 | 14.6 | 172 | 172 | 100 |
|  | MARBLES | 38.2 | 15 | 36.9 | 19 | 238 | 238 | 100 |
| Thallium | EARLI | 0.129 | 0.0736 | 0.114 | 0.077 | 167 | 172 | 97.1 |
|  | MARBLES | 0.122 | 0.0613 | 0.115 | 0.0798 | 220 | 238 | 92.4 |
| Tin | EARLI | 0.574 | 0.894 | 0.303 | 0.395 | 123 | 172 | 71.5 |
|  | MARBLES | 0.579 | 0.69 | 0.354 | 0.493 | 172 | 238 | 72.3 |
| Tungsten | EARLI | 0.0878 | 0.111 | 0.0561 | 0.0931 | 10 | 172 | 5.8 |
|  | MARBLES | 0.143 | 0.232 | 0.103 | 0.153 | 27 | 238 | 11.3 |
| Uranium | EARLI | -0.00802 | 0.0222 | -0.00249 | 0.017 | 12 | 172 | 7 |
|  | MARBLES | 0.00055 | 0.0237 | -0.00091 | 0.0135 | 27 | 238 | 11.3 |
| Vanadium | EARLI | 0.229 | 0.203 | 0.168 | 0.211 | 12 | 172 | 7 |
|  | MARBLES | 0.362 | 0.394 | 0.277 | 0.204 | 6 | 238 | 2.5 |
| Zinc | EARLI | 331 | 203 | 293 | 284 | 171 | 172 | 99.4 |
|  | MARBLES | 334 | 222 | 298 | 260 | 238 | 238 | 100 |

*Acronyms: Early Autism Risk Longitudinal Investigation (EARLI), Markers of Autism Risk in Babies-Learning Early Signs (MARBLES), limit of detection (LOD)*

**Supplemental Table 5**. Maternal and child characteristics of participants with measures of metal exposure in urine from later pregnancy. Data are split by cohort and comparted by neurodevelopmental status of the sibling. Distributions of categorical variables are compared with a chi-square test and continuous variables are compared with an ANOVA test.

| **EARLI cohort** | **Typically developing** | **Non-typically developing** | **Autism spectrum disorder** | **P** |
| --- | --- | --- | --- | --- |
|  | ***N=65*** | ***N=75*** | ***N=31*** |  |
| Maternal Education |  |  |  | 0.016 |
| College Degree | 46 (70.8%) | 40 (53.3%) | 13 (41.9%) |  |
| No Degree | 19 (29.2%) | 35 (46.7%) | 18 (58.1%) |  |
| Maternal Age (years) | 35.0 (4.82) | 33.0 (4.73) | 34.3 (3.86) | 0.038 |
| Infant Sex |  |  |  | 0.004 |
| Female | 38 (58.5%) | 39 (52.0%) | 7 (22.6%) |  |
| Male | 27 (41.5%) | 36 (48.0%) | 24 (77.4%) |  |
| Infant Gestational Age at Sample Collection | 32.8 (3.30) | 32.4 (3.52) | 32.5 (3.99) | 0.787 |
| Infant Gestational Age at Birth (weeks) | 39.5 (1.41) | 39.4 (1.48) | 39.2 (1.48) | 0.496 |
| **MARBLES cohort** | **Typically developing** | **Non-typically developing** | **Autism spectrum disorder** | **P** |
|  | ***N=146*** | ***N=34*** | ***N=51*** |  |
| Maternal Education |  |  |  | 0.124 |
| College Degree | 82 (56.2%) | 15 (44.1%) | 21 (41.2%) |  |
| No Degree | 64 (43.8%) | 19 (55.9%) | 30 (58.8%) |  |
| Maternal Age (years) | 34.0 (4.79) | 33.7 (4.68) | 34.4 (5.40) | 0.806 |
| Infant Sex |  |  |  | 0.167 |
| Female | 71 (48.6%) | 15 (44.1%) | 17 (33.3%) |  |
| Male | 75 (51.4%) | 19 (55.9%) | 34 (66.7%) |  |
| Infant Gestational Age at Sample Collection | 31.4 (3.07) | 31.3 (3.01) | 31.5 (3.36) | 0.957 |
| Infant Gestational Age at Birth (weeks) | 38.9 (1.28) | 39.2 (1.32) | 39.4 (0.99) | 0.021 |

*Acronyms: Early Autism Risk Longitudinal Investigation (EARLI), Markers of Autism Risk in Babies-Learning Early Signs (MARBLES)*

**Supplemental Table 6.** Among participants with two urine metals measures (early and late pregnancy), cross timepoint Spearman correlations for each metal concentration, stratified by cohort (EARLI, MARBLES).

|  | **EARLI (n=165)** | **MARBLES (n=152)** |
| --- | --- | --- |
| Antimony | 0.29 | 0.15 |
| Arsenic | 0.39 | 0.38 |
| Barium | 0.42 | 0.18 |
| Beryllium | 0.07 | 0.20 |
| Cadmium | 0.48 | 0.42 |
| Cesium | 0.42 | 0.42 |
| Chromium | 0.67 | 0.38 |
| Cobalt | 0.28 | 0.24 |
| Copper | 0.33 | 0.39 |
| Lead | 0.49 | 0.26 |
| Manganese | 0.21 | 0.12 |
| Mercury | 0.48 | 0.63 |
| Molybdenum | 0.26 | 0.19 |
| Nickel | 0.29 | 0.18 |
| Platinum | 0.38 | 0.05 |
| Selenium | 0.37 | 0.39 |
| Thallium | 0.39 | 0.33 |
| Tin | 0.57 | 0.59 |
| Tungsten | 0.22 | 0.33 |
| Uranium | 0.45 | 0.20 |
| Vanadium | 0.34 | 0.43 |
| Zinc | 0.53 | 0.47 |

*Acronyms: Early Autism Risk Longitudinal Investigation (EARLI), Markers of Autism Risk in Babies-Learning Early Signs (MARBLES)*

**Supplemental Table 7.** Maternal and child characteristics of participants in the sensitivity analytic sample with measures of metal exposure in blood from pregnancy in the Early Autism Risk Longitudinal Investigation (EARLI) cohort. Data are comparted by neurodevelopmental status of the sibling. Distributions of categorical variables are compared with a chi-square test and continuous variables are compared with an ANOVA test.

|  | **Typically developing** | **Non-typically developing** | **Autism spectrum disorder** | **P-value** |
| --- | --- | --- | --- | --- |
|  | ***N=41*** | ***N=32*** | ***N=19*** |  |
| Maternal Education |  |  |  | 0.175 |
| College Degree | 28 (68.3%) | 16 (50.0%) | 9 (47.4%) |  |
| No Degree | 13 (31.7%) | 16 (50.0%) | 10 (52.6%) |  |
| Maternal Age (years) | 34.4 (4.99) | 33.0 (4.73) | 33.5 (3.95) | 0.446 |
| Infant Sex |  |  |  | 0.001 |
| Female | 24 (58.5%) | 20 (62.5%) | 3 (15.8%) |  |
| Male | 17 (41.5%) | 12 (37.5%) | 16 (84.2%) |  |
| Infant Gestational Age at Birth (weeks) | 39.5 (1.51) | 39.3 (1.69) | 39.1 (1.68) | 0.651 |

**Supplementary Table 8.** Batch adjusted risk ratios for the associations between maternal urinary metal concentrations measured during pregnancy and risk of autism spectrum disorder, relative to typically developing. Log binomial models were adjusted for batch, gestational age, child sex, maternal age, and maternal education. Four metals were modeled categorically (above versus below the limit of detection) and the remaining metals were log_2_ transformed and modeled continuously.

|  | | **EARLI** | | | **MARBLES** | | | **Meta-Analysis** | | | |
| --- | --- | --- | --- | --- | --- | --- | --- | --- | --- | --- | --- |
| **Metal** | **Time** | **RR** | **CI** | **P** | **RR** | **CI** | **P** | **RR** | **CI** | **P** | **FDR** |
| **Modeled categorically** | | | | | | | | | | | |
| Antimony | Early** | 1.70 | (0.92,3.16) | 0.091 | 1.46 | (0.76,2.80) | 0.25 | 1.58 | (1.01,2.48) | 0.044 | 0.37 |
|  | Late | 1.14 | (0.62,2.10) | 0.68 | 1.29 | (0.72,2.34) | 0.39 | 1.22 | (0.79,1.86) | 0.37 | 0.83 |
| Cadmium | Early** | 1.91 | (1.06,3.46) | 0.032 | 1.68 | (0.98,2.87) | 0.06 | 1.78 | (1.19,2.65) | 0.005 | 0.078 |
|  | Late | 1.59 | (0.93,2.73) | 0.091 | 0.91 | (0.52,1.60) | 0.75 | 1.22 | (0.83,1.80) | 0.32 | 0.83 |
| Chromium | Early | 1.18 | (0.65,2.13) | 0.59 | 1.19 | (0.61,2.32) | 0.60 | 1.19 | (0.76,1.84) | 0.45 | 0.69 |
|  | Late | 1.41 | (0.80,2.49) | 0.23 | 0.90 | (0.41,1.99) | 0.79 | 1.21 | (0.76,1.92) | 0.42 | 0.83 |
| Lead | Early | 1.69 | (0.89,3.20) | 0.11 | 1.05 | (0.60,1.86) | 0.86 | 1.30 | (0.85,1.98) | 0.23 | 0.51 |
|  | Late | 1.62 | (0.86,3.05) | 0.14 | 1.04 | (0.65,1.67) | 0.87 | 1.22 | (0.83,1.78) | 0.31 | 0.83 |
| Modeled continuously | | | | | | | | | | | |
| Arsenic | Early** | 0.83 | (0.75,0.91) | <0.001 | 0.97 | (0.76,1.23) | 0.81 | 0.85 | (0.77,0.93) | <0.001 | 0.009 |
|  | Late | 1.16 | (0.91,1.48) | 0.22 | 1.08 | (0.88,1.33) | 0.48 | 1.11 | (0.95,1.30) | 0.18 | 0.68 |
| Barium | Early | 1.13 | (0.88,1.47) | 0.34 | 1.07 | (0.84,1.36) | 0.58 | 1.10 | (0.92,1.31) | 0.29 | 0.57 |
|  | Late | 0.95 | (0.78,1.16) | 0.64 | 0.98 | (0.83,1.16) | 0.81 | 0.97 | (0.85,1.10) | 0.63 | 0.83 |
| Cesium | Early | 1.88 | (0.66,5.39) | 0.24 | 1.53 | (0.70,3.35) | 0.29 | 1.65 | (0.88,3.09) | 0.12 | 0.51 |
|  | Late* | 2.07 | (0.83,5.16) | 0.12 | 1.52 | (0.80,2.89) | 0.20 | 1.69 | (1.00,2.85) | 0.051 | 0.43 |
| Cobalt | Early | 0.87 | (0.55,1.36) | 0.53 | 0.73 | (0.42,1.25) | 0.25 | 0.81 | (0.57,1.14) | 0.22 | 0.51 |
|  | Late | 0.93 | (0.64,1.35) | 0.71 | 1.01 | (0.74,1.37) | 0.97 | 0.98 | (0.77,1.24) | 0.84 | 0.87 |
| Copper | Early | 0.98 | (0.59,1.63) | 0.94 | 1.16 | (0.65,2.06) | 0.61 | 1.06 | (0.72,1.54) | 0.78 | 0.87 |
|  | Late | 3.64 | (1.23,10.79) | 0.02 | 0.94 | (0.72,1.23) | 0.66 | 1.02 | (0.79,1.31) | 0.90 | 0.90 |
| Manganese | Early | 0.75 | (0.44,1.27) | 0.28 | 1.01 | (0.76,1.35) | 0.93 | 0.95 | (0.73,1.22) | 0.66 | 0.83 |
|  | Late | 0.83 | (0.54,1.27) | 0.39 | 1.01 | (0.80,1.29) | 0.90 | 0.97 | (0.79,1.19) | 0.76 | 0.84 |
| Mercury | Early | 1.06 | (0.87,1.30) | 0.56 | 0.79 | (0.58,1.08) | 0.14 | 0.97 | (0.82,1.15) | 0.75 | 0.87 |
|  | Late | 1.14 | (1.02,1.27) | 0.024 | 0.89 | (0.71,1.12) | 0.32 | 1.08 | (0.98,1.20) | 0.11 | 0.58 |
| Molybdenum | Early | 1.85 | (0.82,4.20) | 0.14 | 1.02 | (0.66,1.56) | 0.94 | 1.15 | (0.79,1.69) | 0.46 | 0.69 |
|  | Late | 0.94 | (0.78,1.14) | 0.54 | 1.08 | (0.78,1.48) | 0.64 | 0.98 | (0.83,1.15) | 0.77 | 0.84 |
| Nickel | Early | 1.16 | (0.76,1.78) | 0.49 | 1.04 | (0.57,1.89) | 0.89 | 1.12 | (0.79,1.58) | 0.53 | 0.72 |
|  | Late | 1.09 | (0.72,1.65) | 0.68 | 1.03 | (0.71,1.48) | 0.89 | 1.05 | (0.80,1.39) | 0.70 | 0.83 |
| Selenium | Early | 1.12 | (0.49,2.55) | 0.78 | 0.90 | (0.77,1.06) | 0.21 | 0.91 | (0.78,1.07) | 0.24 | 0.51 |
|  | Late | 2.92 | (0.86,9.96) | 0.087 | 0.91 | (0.75,1.10) | 0.33 | 0.93 | (0.77,1.13) | 0.49 | 0.83 |
| Thallium | Early | 0.91 | (0.56,1.51) | 0.72 | 1.07 | (0.80,1.43) | 0.64 | 1.03 | (0.80,1.32) | 0.82 | 0.87 |
|  | Late | 0.74 | (0.39,1.40) | 0.35 | 1.15 | (0.99,1.33) | 0.072 | 1.12 | (0.97,1.30) | 0.12 | 0.58 |
| Tin | Early | 1.15 | (0.98,1.34) | 0.079 | 0.96 | (0.75,1.23) | 0.76 | 1.09 | (0.96,1.25) | 0.18 | 0.51 |
|  | Late** | 1.17 | (1.03,1.33) | 0.013 | 1.00 | (0.83,1.21) | 1.00 | 1.12 | (1.01,1.24) | 0.04 | 0.43 |
| Zinc | Early | 1.75 | (0.87,3.52) | 0.12 | 0.95 | (0.83,1.08) | 0.42 | 0.97 | (0.85,1.10) | 0.62 | 0.81 |
|  | Late | 1.48 | (0.73,3.01) | 0.28 | 1.08 | (0.81,1.45) | 0.60 | 1.13 | (0.86,1.49) | 0.37 | 0.83 |

*Symbols: ** meta-analysis p-value < 0.05. * meta-analysis p-value < 0.1.*

**Supplementary Table 9.** Batch adjusted risk ratios for the associations between maternal urinary metal concentrations measured during pregnancy and risk of non-typically developing, relative to typically developing. Log binomial models were adjusted for batch, gestational age, child sex, maternal age, and maternal education. Four metals were modeled categorically (above versus below the limit of detection) and the remaining metals were log_2_ transformed and modeled continuously.

|  | | **EARLI** | | | **MARBLES** | | | **Meta-Analysis** | | | |
| --- | --- | --- | --- | --- | --- | --- | --- | --- | --- | --- | --- |
| **Metal** | **Time** | **RR** | **CI** | **P** | **RR** | **CI** | **P** | **RR** | **CI** | **P** | **FDR** |
| **Modeled categorically** | | | | | | | | | | | |
| Antimony | Early | 1.07 | (0.71,1.62) | 0.75 | 0.87 | (0.22,3.43) | 0.85 | 1.05 | (0.71,1.56) | 0.81 | 0.87 |
|  | Late | 0.91 | (0.60,1.37) | 0.64 | 1.73 | (0.87,3.45) | 0.12 | 1.08 | (0.75,1.54) | 0.69 | 0.83 |
| Cadmium | Early** | 1.25 | (0.91,1.72) | 0.17 | 3.10 | (1.33,7.23) | 0.009 | 1.40 | (1.04,1.88) | 0.027 | 0.31 |
|  | Late* | 1.09 | (0.80,1.49) | 0.57 | 2.16 | (1.19,3.93) | 0.011 | 1.26 | (0.96,1.67) | 0.095 | 0.58 |
| Chromium | Early | 0.99 | (0.70,1.40) | 0.97 | 1.33 | (0.48,3.68) | 0.58 | 1.02 | (0.74,1.42) | 0.89 | 0.89 |
|  | Late | 1.01 | (0.67,1.51) | 0.97 | 1.63 | (0.78,3.42) | 0.20 | 1.13 | (0.79,1.61) | 0.51 | 0.83 |
| Lead | Early | 1.11 | (0.80,1.53) | 0.55 | 2.42 | (1.01,5.79) | 0.046 | 1.22 | (0.90,1.66) | 0.21 | 0.51 |
|  | Late | 0.87 | (0.64,1.19) | 0.40 | 1.08 | (0.58,1.99) | 0.81 | 0.91 | (0.69,1.20) | 0.52 | 0.83 |
| Modeled continuously | | | | | | | | | | | |
| Arsenic | Early | 1.10 | (0.95,1.27) | 0.20 | 0.85 | (0.60,1.20) | 0.35 | 1.06 | (0.93,1.21) | 0.40 | 0.69 |
|  | Late | 1.08 | (0.93,1.25) | 0.34 | 0.82 | (0.68,0.98) | 0.032 | 0.96 | (0.86,1.08) | 0.53 | 0.83 |
| Barium | Early | 1.01 | (0.89,1.15) | 0.85 | 1.05 | (0.71,1.54) | 0.82 | 1.02 | (0.90,1.15) | 0.81 | 0.87 |
|  | Late | 0.97 | (0.87,1.09) | 0.64 | 0.96 | (0.77,1.20) | 0.73 | 0.97 | (0.88,1.07) | 0.56 | 0.83 |
| Cesium | Early* | 1.64 | (0.90,2.97) | 0.10 | 1.42 | (0.53,3.78) | 0.48 | 1.58 | (0.95,2.62) | 0.08 | 0.45 |
|  | Late | 1.10 | (0.82,1.46) | 0.53 | 1.06 | (0.55,2.02) | 0.87 | 1.09 | (0.84,1.42) | 0.52 | 0.83 |
| Cobalt | Early | 1.08 | (0.90,1.30) | 0.42 | 1.03 | (0.51,2.09) | 0.94 | 1.07 | (0.90,1.28) | 0.43 | 0.69 |
|  | Late | 0.90 | (0.72,1.13) | 0.38 | 0.77 | (0.51,1.18) | 0.23 | 0.87 | (0.71,1.07) | 0.18 | 0.68 |
| Copper | Early | 1.44 | (0.75,2.77) | 0.27 | 1.48 | (0.58,3.78) | 0.41 | 1.45 | (0.85,2.49) | 0.17 | 0.51 |
|  | Late | 1.06 | (0.73,1.54) | 0.77 | 0.87 | (0.64,1.18) | 0.37 | 0.94 | (0.74,1.19) | 0.61 | 0.83 |
| Manganese | Early | 0.92 | (0.73,1.17) | 0.52 | 0.97 | (0.61,1.55) | 0.90 | 0.93 | (0.75,1.16) | 0.53 | 0.72 |
|  | Late | 0.91 | (0.72,1.16) | 0.45 | 1.05 | (0.79,1.40) | 0.74 | 0.97 | (0.80,1.16) | 0.71 | 0.83 |
| Mercury | Early | 1.06 | (0.99,1.14) | 0.11 | 0.75 | (0.48,1.15) | 0.19 | 1.05 | (0.98,1.13) | 0.18 | 0.51 |
|  | Late | 1.05 | (0.97,1.14) | 0.25 | 0.81 | (0.59,1.11) | 0.19 | 1.03 | (0.95,1.12) | 0.43 | 0.83 |
| Molybdenum | Early | 1.48 | (0.86,2.56) | 0.16 | 1.40 | (0.62,3.16) | 0.42 | 1.45 | (0.92,2.29) | 0.11 | 0.51 |
|  | Late | 1.04 | (0.82,1.32) | 0.75 | 0.92 | (0.72,1.17) | 0.49 | 0.98 | (0.82,1.16) | 0.80 | 0.85 |
| Nickel | Early* | 1.52 | (1.04,2.22) | 0.029 | 0.94 | (0.44,2.02) | 0.88 | 1.39 | (0.99,1.94) | 0.059 | 0.40 |
|  | Late | 1.22 | (0.90,1.65) | 0.20 | 0.74 | (0.59,0.94) | 0.014 | 0.90 | (0.74,1.08) | 0.25 | 0.83 |
| Selenium | Early | 1.25 | (0.69,2.28) | 0.47 | 1.10 | (0.51,2.34) | 0.81 | 1.19 | (0.74,1.90) | 0.47 | 0.69 |
|  | Late** | 1.32 | (0.62,2.83) | 0.47 | 0.85 | (0.81,0.90) | <0.001 | 0.85 | (0.81,0.90) | <0.001 | <0.001 |
| Thallium | Early | 0.75 | (0.44,1.30) | 0.31 | 0.81 | (0.42,1.57) | 0.53 | 0.78 | (0.51,1.18) | 0.24 | 0.51 |
|  | Late | 1.04 | (0.91,1.18) | 0.60 | 0.98 | (0.60,1.58) | 0.92 | 1.03 | (0.91,1.17) | 0.63 | 0.83 |
| Tin | Early | 1.06 | (0.97,1.16) | 0.22 | 0.79 | (0.49,1.27) | 0.33 | 1.05 | (0.96,1.15) | 0.30 | 0.57 |
|  | Late | 1.04 | (0.95,1.14) | 0.42 | 0.83 | (0.61,1.13) | 0.24 | 1.02 | (0.93,1.11) | 0.66 | 0.83 |
| Zinc | Early | 0.99 | (0.86,1.16) | 0.95 | 0.96 | (0.67,1.38) | 0.84 | 0.99 | (0.86,1.14) | 0.89 | 0.89 |
|  | Late** | 0.97 | (0.95,0.98) | <0.001 | 0.92 | (0.82,1.03) | 0.14 | 0.97 | (0.95,0.98) | <0.001 | <0.001 |

*Symbols: ** meta-analysis p-value < 0.05. * meta-analysis p-value < 0.1.*

**Supplementary Table 10.** Adjusted odds ratios for the associations between maternal urinary metal concentrations measured during pregnancy and odds of autism spectrum disorder, relative to typically developing. Logistic regression models were adjusted for gestational age, child sex, maternal age, and maternal education. Four metals were modeled categorically (above versus below the limit of detection) and the remaining metals were log_2_ transformed and modeled continuously.

|  | | **EARLI** | | | **MARBLES** | | | **Meta-Analysis** | | | |
| --- | --- | --- | --- | --- | --- | --- | --- | --- | --- | --- | --- |
| **Metal** | **Time** | **OR** | **CI** | **P** | **OR** | **CI** | **P** | **OR** | **CI** | **P** | **FDR** |
| **Modeled categorically** | | | | | | | | | | | |
| Antimony | Early | 1.19 | (0.37,3.80) | 0.77 | 1.39 | (0.49,3.88) | 0.54 | 1.30 | (0.6,2.8) | 0.51 | 0.71 |
|  | Late | 1.15 | (0.37,3.61) | 0.81 | 1.31 | (0.54,3.19) | 0.55 | 1.25 | (0.62,2.52) | 0.53 | 0.97 |
| Cadmium | Early** | 3.29 | (1.11,9.78) | 0.032 | 2.16 | (0.94,4.98) | 0.071 | 2.52 | (1.3,4.9) | 0.006 | 0.11 |
|  | Late | 2.31 | (0.81,6.58) | 0.12 | 0.95 | (0.44,2.06) | 0.90 | 1.30 | (0.7,2.43) | 0.4 | 0.97 |
| Chromium | Early | 1.76 | (0.55,5.61) | 0.34 | 1.56 | (0.57,4.21) | 0.38 | 1.64 | (0.77,3.49) | 0.20 | 0.55 |
|  | Late | 2.04 | (0.60,6.97) | 0.25 | 0.89 | (0.30,2.64) | 0.84 | 1.28 | (0.57,2.9) | 0.55 | 0.97 |
| Lead | Early | 2.71 | (0.83,8.83) | 0.099 | 1.13 | (0.49,2.58) | 0.78 | 1.50 | (0.76,2.97) | 0.24 | 0.55 |
|  | Late | 2.46 | (0.81,7.46) | 0.11 | 1.11 | (0.57,2.17) | 0.75 | 1.37 | (0.77,2.43) | 0.28 | 0.97 |
| Modeled continuously | | | | | | | | | | | |
| Arsenic | Early | 0.58 | (0.35,0.96) | 0.035 | 1.02 | (0.70,1.46) | 0.93 | 0.84 | (0.62,1.13) | 0.24 | 0.55 |
|  | Late | 1.26 | (0.9,1.76) | 0.18 | 1.13 | (0.87,1.47) | 0.36 | 1.18 | (0.96,1.45) | 0.12 | 0.95 |
| Barium | Early | 1.36 | (0.87,2.12) | 0.18 | 1.10 | (0.78,1.55) | 0.58 | 1.19 | (0.91,1.56) | 0.21 | 0.55 |
|  | Late | 0.97 | (0.67,1.39) | 0.85 | 0.98 | (0.77,1.24) | 0.86 | 0.98 | (0.8,1.19) | 0.80 | 0.97 |
| Cesium | Early** | 3.05 | (0.99,9.36) | 0.051 | 1.70 | (0.75,3.88) | 0.21 | 2.09 | (1.07,4.06) | 0.03 | 0.19 |
|  | Late** | 2.76 | (1.02,7.45) | 0.045 | 1.68 | (0.85,3.31) | 0.14 | 1.97 | (1.12,3.45) | 0.018 | 0.61 |
| Cobalt | Early | 0.73 | (0.36,1.50) | 0.39 | 0.66 | (0.33,1.35) | 0.26 | 0.70 | (0.42,1.15) | 0.16 | 0.55 |
|  | Late | 0.92 | (0.48,1.75) | 0.81 | 1.00 | (0.65,1.54) | 0.99 | 0.97 | (0.68,1.4) | 0.89 | 0.97 |
| Copper | Early | 1.55 | (0.61,3.93) | 0.36 | 1.12 | (0.58,2.16) | 0.74 | 1.25 | (0.73,2.13) | 0.42 | 0.68 |
|  | Late | 3.76 | (1.41,10.01) | 0.008 | 0.89 | (0.58,1.37) | 0.60 | 1.12 | (0.76,1.66) | 0.56 | 0.97 |
| Manganese | Early | 0.92 | (0.52,1.62) | 0.77 | 1.08 | (0.70,1.66) | 0.73 | 1.02 | (0.72,1.44) | 0.92 | 0.93 |
|  | Late | 0.89 | (0.56,1.43) | 0.64 | 1.01 | (0.71,1.42) | 0.97 | 0.96 | (0.73,1.27) | 0.80 | 0.97 |
| Mercury | Early | 1.38 | (0.88,2.14) | 0.16 | 0.78 | (0.55,1.09) | 0.15 | 0.96 | (0.73,1.26) | 0.77 | 0.88 |
|  | Late | 1.43 | (0.88,2.33) | 0.15 | 0.89 | (0.68,1.15) | 0.37 | 0.99 | (0.78,1.24) | 0.91 | 0.97 |
| Molybdenum | Early | 1.56 | (0.82,2.98) | 0.18 | 0.96 | (0.54,1.71) | 0.89 | 1.19 | (0.77,1.83) | 0.43 | 0.68 |
|  | Late | 0.85 | (0.48,1.51) | 0.58 | 1.10 | (0.76,1.59) | 0.61 | 1.02 | (0.75,1.39) | 0.90 | 0.97 |
| Nickel | Early | 1.09 | (0.57,2.07) | 0.80 | 1.06 | (0.47,2.36) | 0.89 | 1.08 | (0.65,1.78) | 0.78 | 0.88 |
|  | Late | 1.13 | (0.58,2.22) | 0.72 | 1.01 | (0.62,1.66) | 0.96 | 1.05 | (0.71,1.57) | 0.80 | 0.97 |
| Selenium | Early | 0.90 | (0.33,2.42) | 0.83 | 0.82 | (0.44,1.55) | 0.55 | 0.84 | (0.5,1.44) | 0.54 | 0.71 |
|  | Late | 2.20 | (0.79,6.13) | 0.13 | 0.87 | (0.49,1.55) | 0.63 | 1.09 | (0.66,1.8) | 0.74 | 0.97 |
| Thallium | Early | 1.01 | (0.49,2.07) | 0.98 | 1.35 | (0.81,2.26) | 0.26 | 1.22 | (0.8,1.86) | 0.35 | 0.66 |
|  | Late | 0.78 | (0.43,1.41) | 0.41 | 1.46 | (0.93,2.30) | 0.099 | 1.16 | (0.81,1.66) | 0.42 | 0.97 |
| Tin | Early | 1.42 | (0.92,2.20) | 0.11 | 0.97 | (0.69,1.35) | 0.85 | 1.11 | (0.86,1.45) | 0.42 | 0.68 |
|  | Late | 1.48 | (0.99,2.20) | 0.055 | 1.02 | (0.78,1.32) | 0.89 | 1.14 | (0.92,1.42) | 0.25 | 0.97 |
| Zinc | Early | 1.95 | (1.05,3.61) | 0.034 | 0.90 | (0.62,1.32) | 0.60 | 1.12 | (0.81,1.54) | 0.51 | 0.71 |
|  | Late | 1.54 | (0.83,2.88) | 0.17 | 1.11 | (0.81,1.53) | 0.52 | 1.19 | (0.9,1.58) | 0.23 | 0.97 |

*Symbols: ** meta-analysis p-value < 0.05. * meta-analysis p-value < 0.1.*

**Supplementary Table 11.** Adjusted odds ratios for the associations between maternal urinary metal concentrations measured during pregnancy and odds of non-typically developing, relative to typically developing. Logistic regression models were adjusted for gestational age, child sex, maternal age, and maternal education. Four metals were modeled categorically (above versus below the limit of detection) and the remaining metals were log_2_ transformed and modeled continuously.

|  | | **EARLI** | | | **MARBLES** | | | **Meta-Analysis** | | | |
| --- | --- | --- | --- | --- | --- | --- | --- | --- | --- | --- | --- |
| **Metal** | **Time** | **OR** | **CI** | **P** | **OR** | **CI** | **P** | **OR** | **CI** | **P** | **FDR** |
| **Modeled categorically** | | | | | | | | | | | |
| Antimony | Early | 1.49 | (0.67,3.32) | 0.33 | 0.66 | (0.13,3.25) | 0.61 | 1.26 | (0.62,2.58) | 0.53 | 0.71 |
|  | Late | 0.92 | (0.4,2.09) | 0.84 | 1.98 | (0.77,5.09) | 0.16 | 1.28 | (0.69,2.38) | 0.44 | 0.97 |
| Cadmium | Early** | 1.76 | (0.86,3.6) | 0.12 | 4.12 | (1.4,12.1) | 0.01 | 2.29 | (1.26,4.15) | 0.007 | 0.11 |
|  | Late* | 1.24 | (0.61,2.53) | 0.55 | 2.5 | (1.1,5.67) | 0.028 | 1.68 | (0.98,2.88) | 0.059 | 0.95 |
| Chromium | Early | 0.93 | (0.43,2.03) | 0.86 | 1.6 | (0.46,5.65) | 0.46 | 1.08 | (0.56,2.1) | 0.81 | 0.89 |
|  | Late | 1.02 | (0.4,2.62) | 0.97 | 1.7 | (0.6,4.77) | 0.32 | 1.29 | (0.64,2.58) | 0.48 | 0.97 |
| Lead | Early* | 1.27 | (0.61,2.64) | 0.53 | 3.04 | (1.04,8.89) | 0.042 | 1.68 | (0.92,3.07) | 0.094 | 0.46 |
|  | Late | 0.71 | (0.34,1.49) | 0.37 | 1.11 | (0.52,2.39) | 0.79 | 0.88 | (0.52,1.5) | 0.64 | 0.97 |
| Modeled continuously | | | | | | | | | | | |
| Arsenic | Early | 1.20 | (0.95,1.52) | 0.12 | 0.84 | (0.50,1.41) | 0.51 | 1.13 | (0.92,1.4) | 0.25 | 0.55 |
|  | Late | 1.16 | (0.89,1.51) | 0.28 | 0.73 | (0.52,1.03) | 0.078 | 0.98 | (0.79,1.21) | 0.83 | 0.97 |
| Barium | Early | 1.00 | (0.75,1.33) | 0.99 | 1.05 | (0.66,1.66) | 0.85 | 1.01 | (0.79,1.29) | 0.93 | 0.93 |
|  | Late | 0.92 | (0.71,1.20) | 0.54 | 0.95 | (0.72,1.26) | 0.74 | 0.94 | (0.77,1.13) | 0.50 | 0.97 |
| Cesium | Early** | 2.16 | (1.04,4.50) | 0.039 | 1.49 | (0.53,4.18) | 0.45 | 1.91 | (1.05,3.47) | 0.034 | 0.19 |
|  | Late | 1.15 | (0.68,1.95) | 0.60 | 1.07 | (0.49,2.33) | 0.86 | 1.13 | (0.73,1.74) | 0.60 | 0.97 |
| Cobalt | Early | 1.25 | (0.73,2.14) | 0.41 | 1.07 | (0.44,2.58) | 0.88 | 1.20 | (0.76,1.9) | 0.44 | 0.68 |
|  | Late | 0.79 | (0.50,1.24) | 0.30 | 0.72 | (0.43,1.22) | 0.22 | 0.76 | (0.54,1.07) | 0.11 | 0.95 |
| Copper | Early | 1.34 | (0.71,2.54) | 0.37 | 1.37 | (0.55,3.43) | 0.5 | 1.35 | (0.8,2.28) | 0.26 | 0.55 |
|  | Late | 1.01 | (0.58,1.77) | 0.97 | 0.81 | (0.48,1.38) | 0.44 | 0.90 | (0.61,1.32) | 0.60 | 0.97 |
| Manganese | Early | 0.80 | (0.56,1.14) | 0.23 | 1.01 | (0.58,1.74) | 0.98 | 0.86 | (0.64,1.16) | 0.31 | 0.62 |
|  | Late | 0.86 | (0.64,1.16) | 0.31 | 1.07 | (0.73,1.56) | 0.75 | 0.93 | (0.74,1.18) | 0.55 | 0.97 |
| Mercury | Early | 1.11 | (0.83,1.47) | 0.48 | 0.75 | (0.48,1.17) | 0.20 | 0.99 | (0.78,1.26) | 0.93 | 0.93 |
|  | Late | 1.08 | (0.80,1.46) | 0.60 | 0.81 | (0.59,1.11) | 0.18 | 0.94 | (0.76,1.17) | 0.59 | 0.97 |
| Molybdenum | Early** | 1.64 | (1.03,2.61) | 0.038 | 1.34 | (0.61,2.94) | 0.47 | 1.55 | (1.04,2.32) | 0.031 | 0.19 |
|  | Late | 1.09 | (0.73,1.61) | 0.68 | 0.91 | (0.58,1.41) | 0.67 | 1.00 | (0.75,1.35) | 0.98 | 0.98 |
| Nickel | Early** | 2.10 | (1.27,3.46) | 0.0036 | 0.90 | (0.35,2.33) | 0.82 | 1.75 | (1.12,2.72) | 0.013 | 0.15 |
|  | Late | 1.47 | (0.91,2.36) | 0.12 | 0.62 | (0.35,1.09) | 0.098 | 1.03 | (0.71,1.48) | 0.88 | 0.97 |
| Selenium | Early | 1.50 | (0.85,2.64) | 0.16 | 1.15 | (0.52,2.53) | 0.73 | 1.37 | (0.86,2.17) | 0.18 | 0.55 |
|  | Late | 1.43 | (0.71,2.89) | 0.32 | 0.67 | (0.33,1.36) | 0.27 | 0.98 | (0.6,1.62) | 0.95 | 0.98 |
| Thallium | Early | 0.64 | (0.37,1.09) | 0.10 | 0.96 | (0.51,1.78) | 0.89 | 0.76 | (0.5,1.14) | 0.19 | 0.55 |
|  | Late | 1.03 | (0.68,1.56) | 0.87 | 1.02 | (0.64,1.64) | 0.92 | 1.03 | (0.75,1.41) | 0.86 | 0.97 |
| Tin | Early | 1.15 | (0.86,1.53) | 0.35 | 0.81 | (0.50,1.32) | 0.40 | 1.05 | (0.82,1.34) | 0.71 | 0.87 |
|  | Late | 1.08 | (0.84,1.39) | 0.55 | 0.82 | (0.59,1.14) | 0.24 | 0.97 | (0.8,1.19) | 0.80 | 0.97 |
| Zinc | Early | 0.95 | (0.66,1.38) | 0.79 | 0.94 | (0.57,1.57) | 0.82 | 0.95 | (0.7,1.28) | 0.72 | 0.87 |
|  | Late | 0.81 | (0.58,1.12) | 0.21 | 0.86 | (0.60,1.24) | 0.42 | 0.83 | (0.65,1.06) | 0.14 | 0.95 |

*Symbols: ** meta-analysis p-value < 0.05. * meta-analysis p-value < 0.1.*

**Supplemental Figure 1.** Gestation age at urine sample collection, separated by the earlier pregnancy and later pregnancy timepoints in A) EARLI cohort and B) MARBLES cohort.

A. **EARLI**

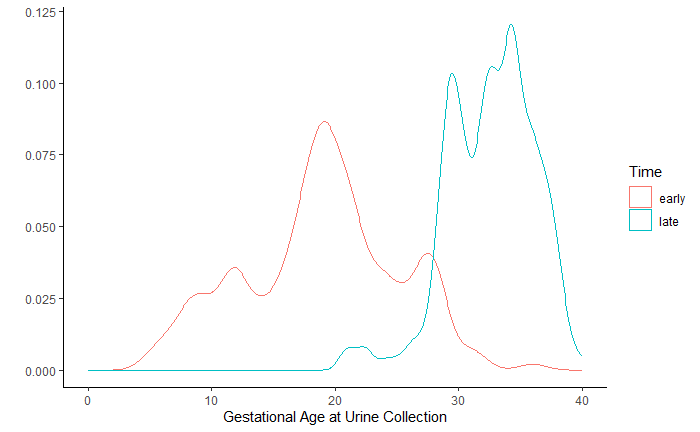

B. **MARBLES**

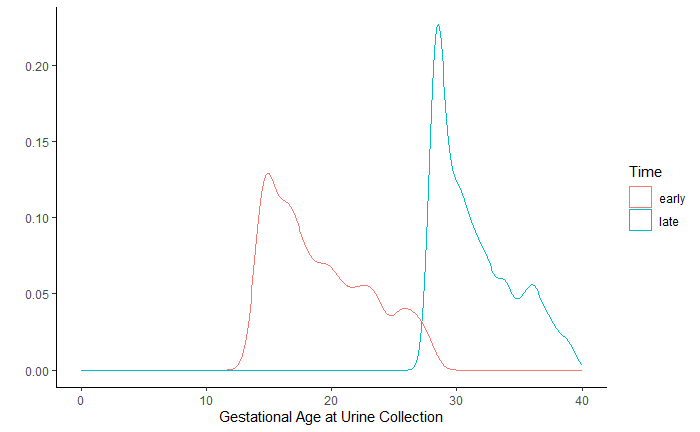

**Supplemental Figure 2.** Spearman correlations of urinary metals concentrations, measured during late pregnancy, stratified by cohort. The upper right triangle shows the EARLI cohort. The lower left triangle shows the MARBLES cohort. Metals are represented by their chemical symbol along the diagonal.

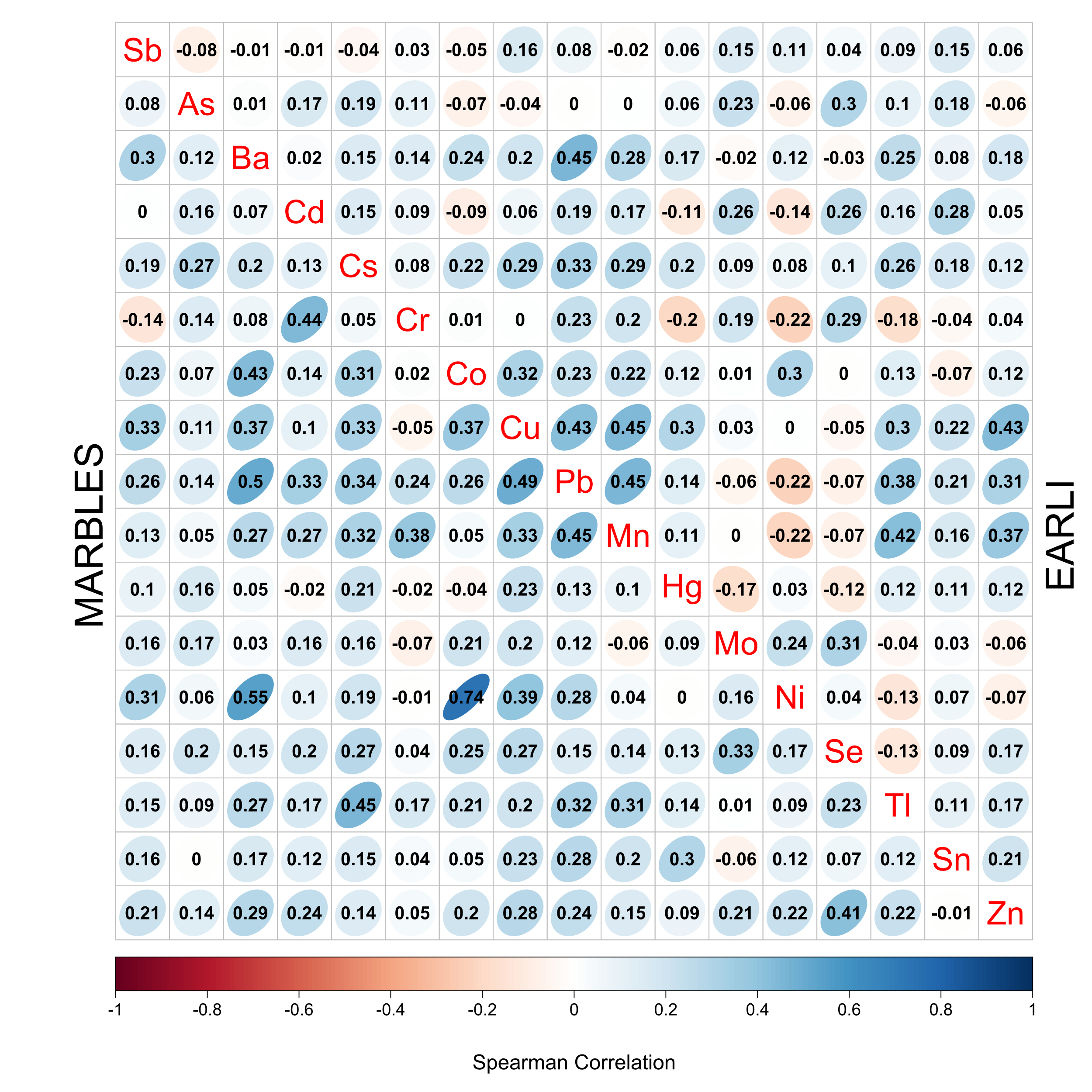

*Acronyms: Early Autism Risk Longitudinal Investigation (EARLI), Markers of Autism Risk in Babies-Learning Early Signs (MARBLES)*

**Supplemental Figure 2**. Adjusted associations between maternal blood metals concentrations and infant neurodevelopmental status in the Early Autism Risk Longitudinal Investigation (EARLI). **A)** Comparison between autism spectrum disorder relative to typically developing. **B)** Comparison between non-typically developing relative to typically developing. Relative risk ratios are reported for a doubling in concentration.

| **A.**  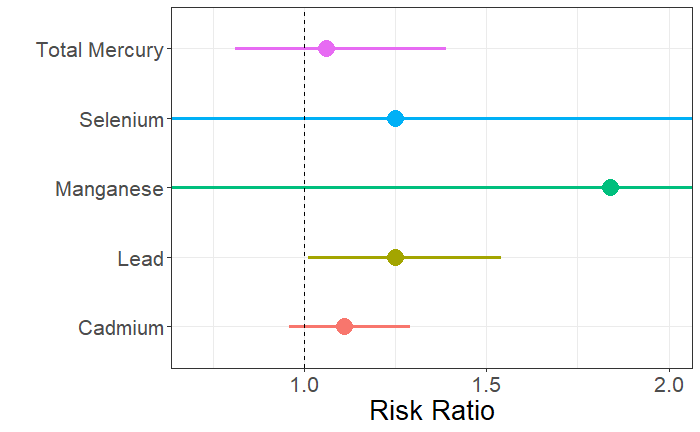 | **B.**  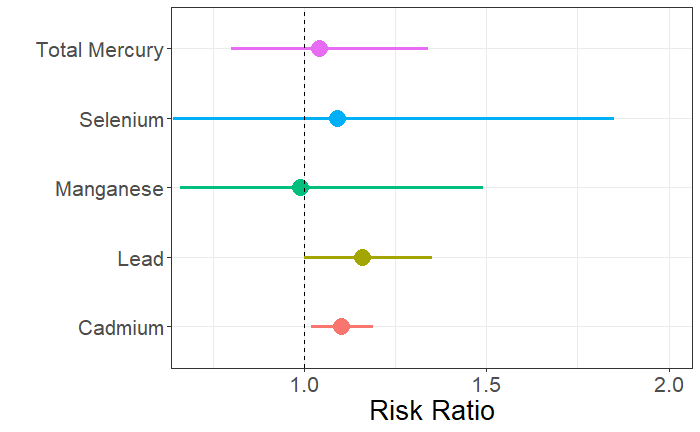 |
| --- | --- |
